# Genome-wide QTL mapping across three tissues highlights several Alzheimer’s and Parkinson’s disease loci potentially acting via DNA methylation

**DOI:** 10.1101/2023.12.22.23300365

**Authors:** Olena Ohlei, Yasmine Sommerer, Valerija Dobricic, Jan Homann, Laura Deecke, Marcel Schilling, David Bartrés-Faz, Gabriele Cattaneo, Sandra Düzel, Anders M. Fjell, Ulman Lindenberger, Álvaro Pascual-Leone, Sanaz Sedghpour Sabet, Cristina Solé-Padullés, Josep M. Tormos, Valentin M. Vetter, Kristine B. Walhovd, Tanja Wesse, Michael Wittig, Andre Franke, Ilja Demuth, Christina M. Lill, Lars Bertram

**Affiliations:** Lübeck Interdisciplinary Platform for Genome Analytics, University of Lübeck, Lübeck, Germany; Institute of Epidemiology and Social Medicine, University of Münster, Münster, Germany; Gene Regulation of Cell Identity, Regenerative Medicine Program, Bellvitge Institute for Biomedical; Research (IDIBELL), L’Hospitalet del Llobregat, Barcelona, Spain; Department of Medicine, Faculty of Medicine and Health Sciences, Institute of Neurosciences, University of Barcelona, Barcelona, Spain; Institut Guttmann, Institut Universitari de Neurorehabilitació adscrit a la UAB, Barcelona, Spain; Departament de Medicina, Universitat Autònoma de Barcelona, Barcelona, Spain; Fundació Institut d’Investigació en Ciències de la Salut Germans Trias i Pujol, Camí de les Escoles, Badalona, Barcelona, Spain; Center for Lifespan Psychology, Max Planck Institute for Human Development, Berlin, Germany; Center for Lifespan Changes in Brain and Cognition, University of Oslo, Oslo, Norway; Department of Radiology and Nuclear Medicine, Oslo University Hospital, Oslo, Norway; Hinda and Arthur Marcus Institute for Aging Research and Deanna and Sidney Wolk Center for Memory Health, Hebrew SeniorLife, Boston, MA, USA; Department of Neurology, Harvard Medical School, Boston, MA, USA; Institute of Clinical Molecular Biology, Christian-Albrechts-University, Kiel, Germany; Biology of Aging Working Group, Department of Endocrinology and Metabolic Diseases, Division of Lipid Metabolism, Charité—Universitätsmedizin Berlin (corporate member of Freie Universität Berlin and Humboldt-Universität zu Berlin), Berlin, Germany; Berlin Institute of Health Center for Regenerative Therapies, Charité—Universitätsmedizin Berlin, Berlin, Germany; Ageing Epidemiology Research Unit (AGE), School of Public Health, Imperial College London, London, UK

**Keywords:** Genome-wide association study (GWAS), methylation quantitative trait locus (meQTL) analysis, Mendelian randomization (MR), colocalization, Alzheimer’s disease (AD), Parkinson’s disease (PD)

## Abstract

DNA methylation (DNAm) is an epigenetic mark with essential roles in disease development and predisposition. Here, we created genome-wide maps of methylation quantitative trait loci (meQTL) in three peripheral tissues and used Mendelian randomization (MR) analyses to assess the potential causal relationships between DNAm and risk for two common neurodegenerative disorders, i.e. Alzheimer’s disease (AD) and Parkinson’s disease (PD). Genome-wide single nucleotide polymorphism (SNP; ∼5.5M sites) and DNAm (∼850K CpG sites) data were generated from whole blood (n=1,058), buccal (n=1,527) and saliva (n=837) specimens. We identified between 11 and 15 million genome-wide significant (p<10^-14^) SNP-CpG associations in each tissue. Combining these meQTL GWAS results with recent AD/PD GWAS summary statistics by MR strongly suggests that the previously described associations between *PSMC3*, *PICALM*, and *TSPAN14* and AD may be founded on differential DNAm in or near these genes. In addition, there is strong, albeit less unequivocal, support for causal links between DNAm at *PRDM7* in AD as well as at *KANSL1/MAPT* in AD and PD. Our study adds valuable insights on AD/PD pathogenesis by combining two high-resolution “omics” domains, and the meQTL data shared along with this publication will allow like-minded analyses in other diseases.

## Introduction

Genome-wide associations studies (GWAS) in Alzheimer’s disease (AD)^1^ and Parkinson’s disease (PD)^2^ continue to identify an increasing number of genetic variants associated with the risk for developing these neurodegenerative disorders. However, the functional mechanisms underlying these associations remain largely unknown. An individual’s predisposition for developing AD or PD results from the complex interplay between genetic and non-genetic factors unfolding their effects over a person’s lifetime^3,4^. Examples of non-genetic factors are lifestyle variables (e.g. smoking, nutritional habits), environmental exposures (e.g. air quality, place of residence), and epigenetic mechanisms (e.g. DNA methylation [DNAm], histone modifications). In the context of complex disease research, epigenetic mechanisms play a particularly interesting role as they lie at the intersection between lifestyle/environment, genetics, and the regulation of gene expression^5–8^. In this context, DNAm is one of the most widely studied epigenetic marks owing to the advent of high-throughput technologies allowing the interrogation of DNAm profiles on a genome-wide scale.

One method to probe for epigenetic effects on disease risk is to conduct epigenome-wide association studies (EWAS). In AD, several such EWAS have been published using both brain^9–13^ and peripheral tissues^14–16^ highlighting a number of CpG sites that show differential DNAm with respect to disease state. Similar EWAS efforts have been completed in PD, e.g. using brain^17^ and blood^18^. Collectively, these studies have led to the initial delineation of DNAm profiles associated with these disorders. Despite this progress, one major caveat of most published EWAS is that cause-effect relationships are difficult to discern, i.e. to distinguish whether the observed differential DNAm patterns contribute to pathogenesis, and as such occur before or early during the disease process (potentially highlighting disease-causing mechanisms) or whether they are the result of the disease process itself (e.g. due to the accumulation of pathologic protein aggregates). One possibility to solve this inference problem is to use genetics as a “common denominator” variable, e.g. in the context of Mendelian Randomization (MR) analyses. In MR, which is a type of instrumental variable analysis, genetic risk variants (e.g. from GWAS) are combined with genetic variants affecting the exposure of interest (here: DNAm), so called methylation quantitative trait loci (meQTLs), allowing to draw direct inferences on a causal relationship between the two. If interpreted carefully^19,20^, MR can effectively shed new light on the “causality uncertainty” in EWAS.

A prerequisite for meQTL-based MR analyses is the availability of meQTL GWAS data in the tissues of interest. For instance, meQTL GWAS have been performed in blood^21,22^, brain^23–25^, buccal^26^, and saliva samples^27,28^, although these initial reports used comparatively low-resolution DNAm microarrays (with n<500K CpG markers). One of the currently largest meQTL GWAS in terms of sample size was recently published for ∼7,000 blood samples from two different ethnic descent groups, European and Asian^22^. DNAm profiling in that study was based on the Illumina Infinium HumanMethylation450 BeadChip (450K), capturing ∼450K CpG sites. The study identified approximately 11.2 million genome-wide significant SNP-CpG pairs, of which ∼50% showed high cross-tissue correspondence. Another noteworthy meQTL study was recently published by the Genotype-Tissue Expression (GTEx; www.gtexportal.org) project which examined nine tissues from ∼400 donors in parallel (breast, kidney, colon, lung, muscle, ovary, prostate, testis and whole blood)^29^. While considerably smaller in sample size than ref. 22, the GTEx team used the Illumina successor array, (Infinium MethylationEPIC [EPIC], containing nearly 850K CpGs)^29^. Neither study performed systematic MR analyses to quantify the impact of DNAm on disease risk.

To close these gaps, we have created extensive genome-wide meQTL maps for three peripheral tissues (blood, buccals, and saliva) using the currently highest-resolution commercial DNAm microarray (EPIC) in sample sizes ranging from n=837 (saliva) to n=1,527 (buccals). Each of these meQTL GWAS identified between 11 and 15 million genome-wide significant (p < 10^-14^) SNP-CpG pairs, a large fraction showing high cross-tissue correspondence. Next, we combined these novel meQTL GWAS results with recent risk GWAS for AD^1^ (n= 1,474,097) and PD^2^ (n= 788,989) using various different MR and colocalization analysis paradigms to assess whether and which of the hitherto reported GWAS signals might unfold their effects by affecting DNAm. Our novel results strongly suggest that the GWAS-based risk associations between up to five known AD/PD GWAS loci may at least partially be due to differential methylation. The complete and novel meQTL GWAS results, which provide the backbone of our study, are made freely available (URL: https://doi.org/10.5281/zenodo.10410506) for use in like-minded analyses on different phenotypes.

## Results

### Methylation quantitative trait locus (meQTL) genome-wide association studies (GWAS) and independent replication analyses

For each of the three available tissues, blood (n=1,058), buccals (n=1,527) and saliva (n=837), we performed meQTL GWAS analyses testing approximately 5.5 million common (minor allele frequency [MAF] ≥0.05) SNPs for association with approximately 750,000 CpG probes after QC (**Figure 1**). In total, this procedure resulted in over 12 trillion statistical tests, of which approximately 1.7 trillion were independent, resulting in a conservative study-wide α-level of 1×10^-14^ (Methods). Using this threshold, we identified between 11 and 15 million genome-wide significant SNP-CpG pairs in each dataset (**Figure 2**). In blood, approximately 92% of meQTL were detected in *cis* (i.e. SNP-CpG distance within ±1MB on the same chromosome), whereas 4% meQTL were in long-range *cis* (lr-cis, i.e. SNP-CpG distance >1Mb but located on the same chromosome), and 4% meQTL were in *trans* (i.e. SNP and CpG sites were located on different chromosomes). Comparable numbers of meQTLs were identified in GWAS analyses of buccal and saliva samples (**Table 1**; Supplementary Tables S1-S3). To the best of our knowledge, our study comprises the largest meQTL GWAS available for these latter two tissues to date.

**Figure 1.**
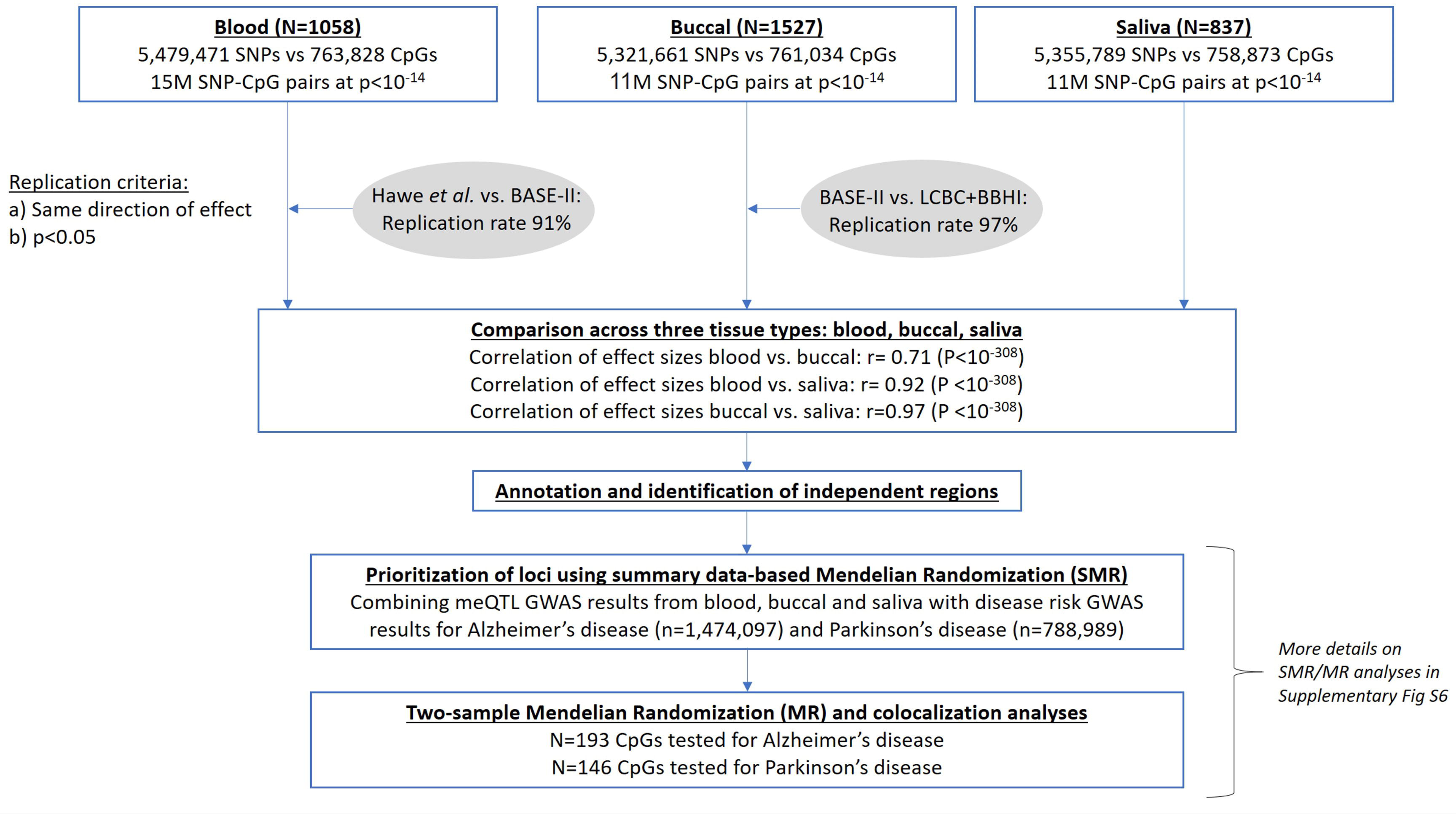
Flowchart of meQTL study design and analysis strategies applied in this work. More details on the SMR/MR analyses can be found in Supplementary Figure S6.

**Figure 2.**
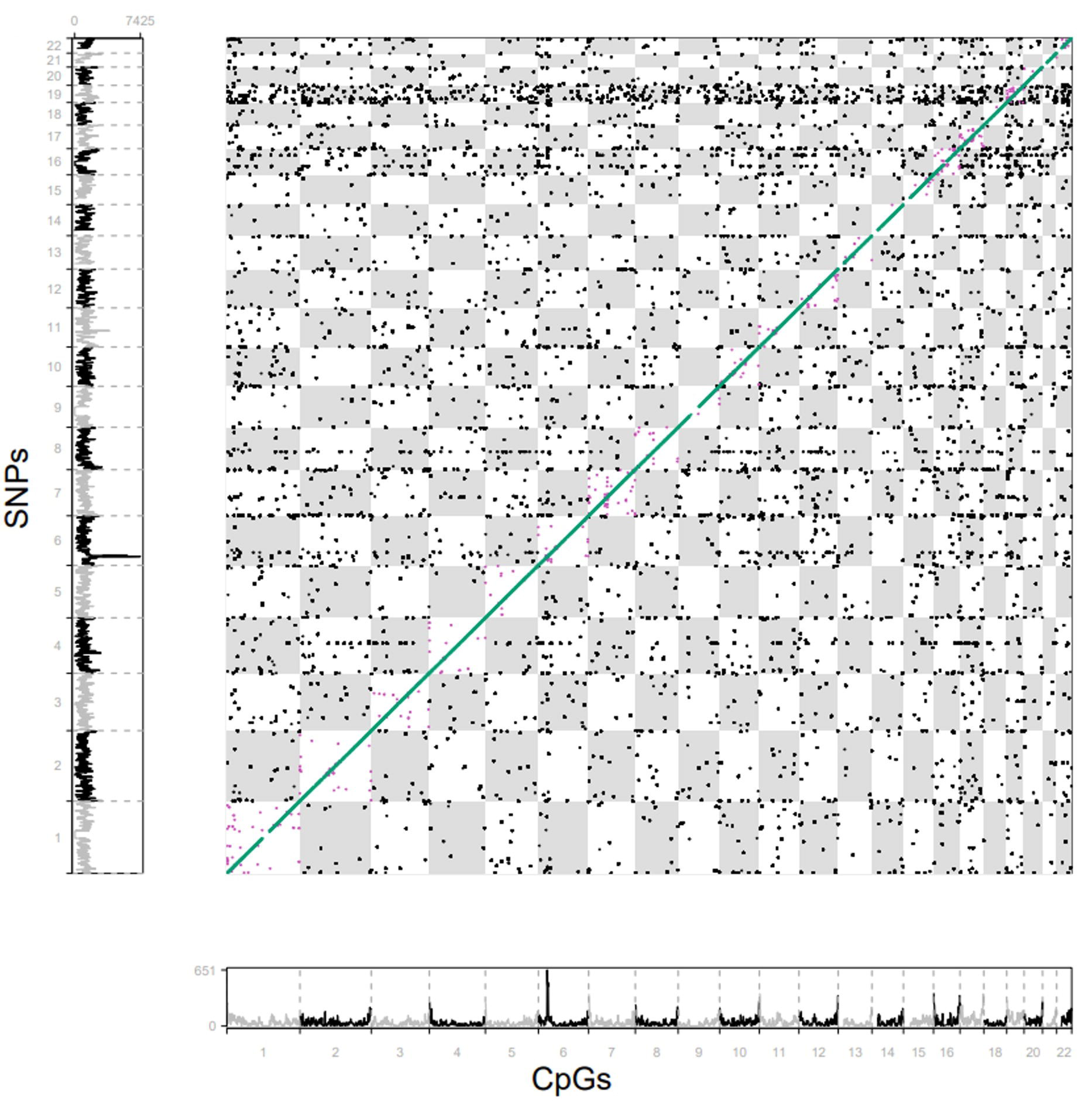
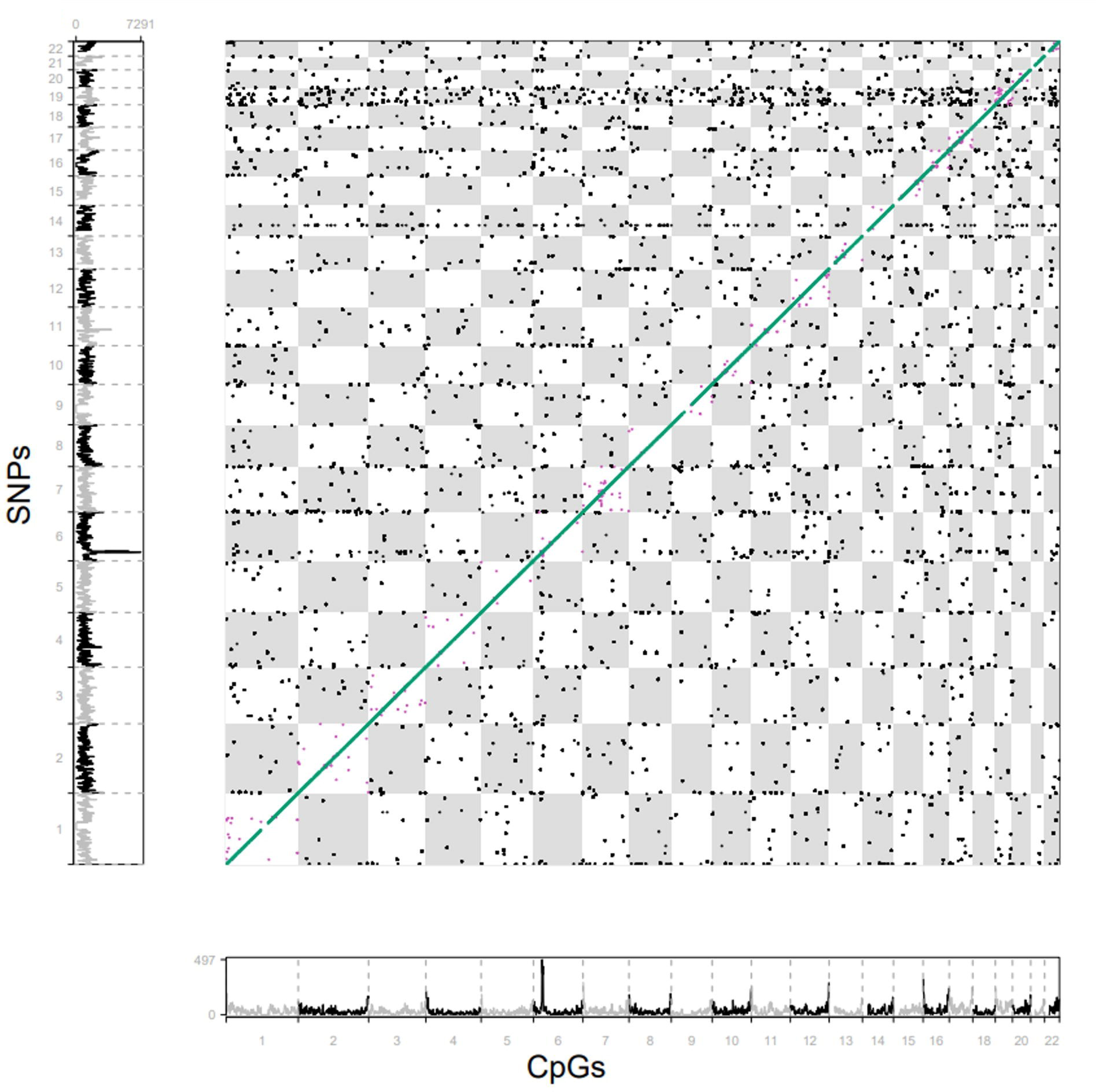
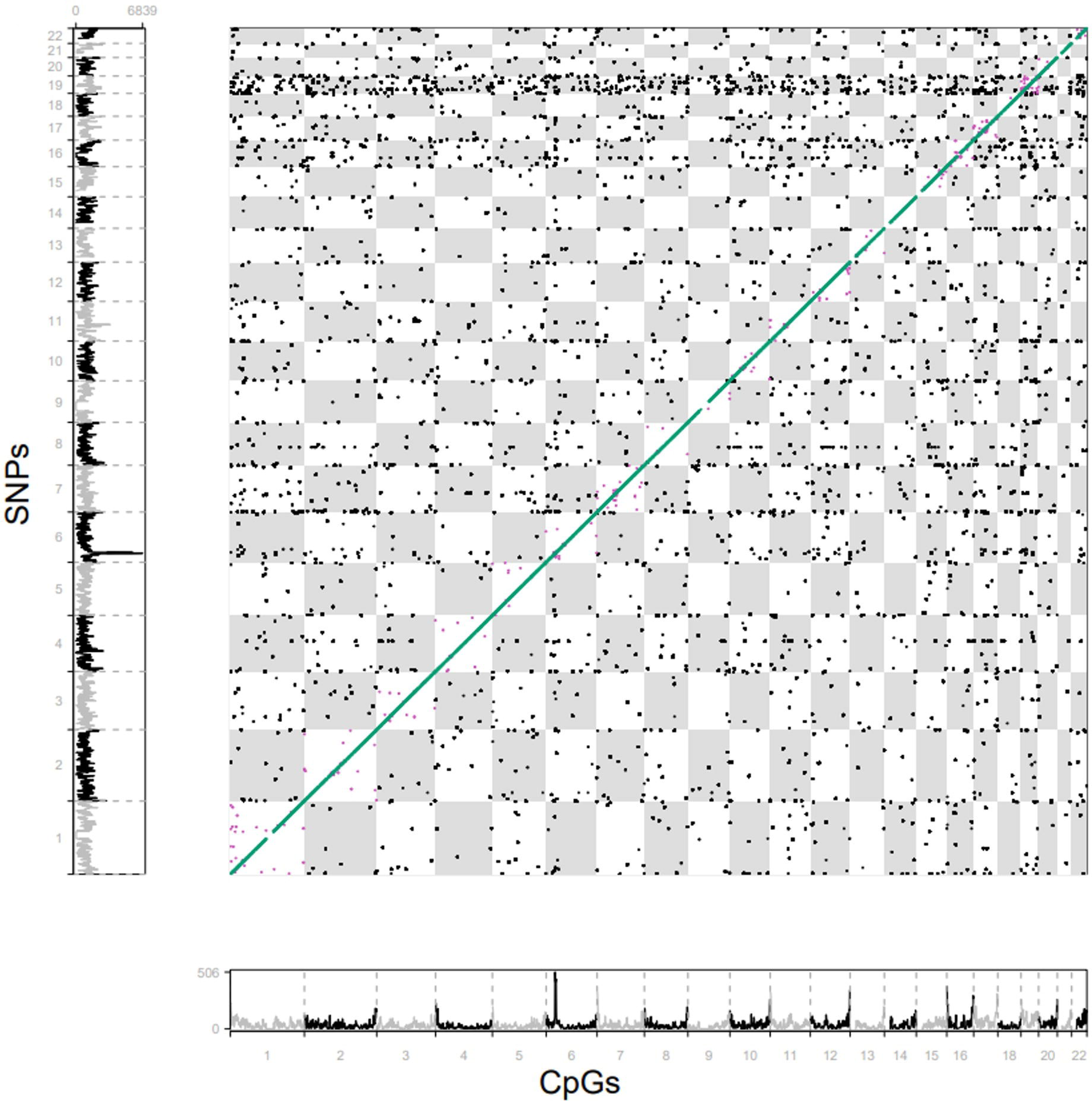
Chessboard plots for study-wide significant SNP-CpG associations results in all three tissue types analyzed here: blood, buccal mucosa and saliva. A. SNP-CpG pairwise associations in blood dataset (i.e. BASE-II; n=1,058). B. SNP-CpG pairwise associations in available buccal datasets (i.e. BASE-II, LCBC and BBHI; n=1,527). C. SNP-CpG pairwise associations in saliva dataset (i.e. LCBC; n=837). Each dot represents a SNP-CpG pair that has exceeded the study-wide significance level (P<10^-14^; Methods). CpG positions are shown on the x-axis, and SNP positions are shown on the y-axis. CpG position and CpG density (#CpGs/Mb) are provided on the x axis, while SNP position and SNP density (#CpGs/Mb) are provided on the y axis. SNP-CpG pairs are coded according to their genomic distance: *cis* = for pairs within 1 Mb (green markers; appear as a diagonal line); long-range *cis* = for pairs on the same chromosome but >1Mb apart (purple markers); *trans* = for pairs located on different chromosomes (black markers).

**Table 1.** Number of significant (p < 10^-14^) meQTL results across tissue types.

| meQTL | Blood<br>(# CpGs/# SNPs; %) | Buccal<br>(# CpGs/# SNPs; %) | Saliva<br>(# CpGs/# SNPs; %) |
| --- | --- | --- | --- |
| *cis* | 13,861,999<br>(118,955/2,955,700; 0.92) | 10,610,037<br>(92,694/2,792,056; 0.92) | 10,683,104<br>(100,233/2,648,648; 0.93) |
| long-range *cis* | 606,035<br>(2,066/102,826; 0.04) | 531,305<br>(1,645/94,094; 0.05) | 479,377<br>(1,714/89,074; 0.04) |
| *trans* | 551,691<br>(4,954/287,333; 0.04) | 347,081<br>(3,156/208,584; 0.03) | 374,550<br>(3,504/199,877; 0.03) |
| total | 15,019,725<br>(123,703/3,010,228; *n.a.*) | 11,488,423<br>(95,791/2,837,984; *n.a.*) | 11,537,03<br>(103,668/2,690,318; *n.a.*) |

For buccal swab specimen, we had two independent datasets (**Figure 1**, Supplementary Figure S2) available allowing us to assess the degree of replication for meQTL associations within that particular tissue. These analyses revealed a very high (∼94%) degree of replication of SNP-CpG pairs showing genome-wide significance in BASE-II samples (n = 837) when assessed in the combined BBHI-LCBC-buccal (n=690) dataset. For this purpose, “replication” was assumed for SNP-CpG pairs showing the same direction of effect with at least nominal significance (i.e. p<0.05; Methods) as suggested previously^22^. To assess replication in blood, we compared our meQTL results to the findings recently reported by Hawe et al.^22^. Of the 11,165,559 genome-wide significant “cosmopolitan” meQTLs showing both ancestry-specific replication in samples from Europe and Asia^22^, 7,612,751 were also tested in BASE-II blood samples; of these 7,405,579 (∼97%) showed consistent effect directions with at least nominal significance (p<0.05). Furthermore, we found a highly significant correlation of effect size estimates between significant (p<10^-14^) cosmopolitan meQTLs results from Hawe at al.^22^ and our analyses (r= 0.96, p<2.2×10^-16^). While no independent dataset of sufficient size was available to assess replication of meQTL effects in saliva, overall, these findings demonstrate that our meQTL results are highly robust and, for blood, are in good agreement with the literature.

### Comparison of meQTL findings across three tissues: blood, buccals, saliva

Next, we addressed the question as to how stable meQTL effects were across tissue types by estimating cross-tissue correspondence rates (using the same criteria defining replication outlined above). Since blood and buccal tissue data were partially obtained from the same individuals (i.e. participants of the BASE-II study; **Figure 1**) we used the independent blood meQTL results recently reported by Hawe et al.^22^ to compare replication and correlation of effect direction with the other tissues (**Figure 3B-D**). Overall, we observe high degrees of cross-tissue correspondence for *cis* SNP-CpG pairs ranging from 71 to 94% across all three tissue types. The highest correspondence (94/96/97%) was seen when comparing *cis*/lr-*cis*/*trans* SNP-CpG pairs in buccal vs. saliva specimen (**Figure 3D**), while *cis*/lr-*cis*/*trans* SNP-CpG pairs in blood vs. buccals (**Figure 3B**) showed the lowest correspondence rates (71/74/74%). In contrast, the strongest correlation of effect sizes (r=0.92/0.92/0.90) was observed when comparing *cis*/lr-*cis*/*trans* SNP-CpG pairs in blood vs. saliva specimen (**Figure 3C**), while *cis*/lr-*cis*/*trans* SNP-CpG pairs in blood vs. buccals (**Figure 3B**) showed the weakest correlations (r=0.71/0.80/0.82).

**Figure 3.**
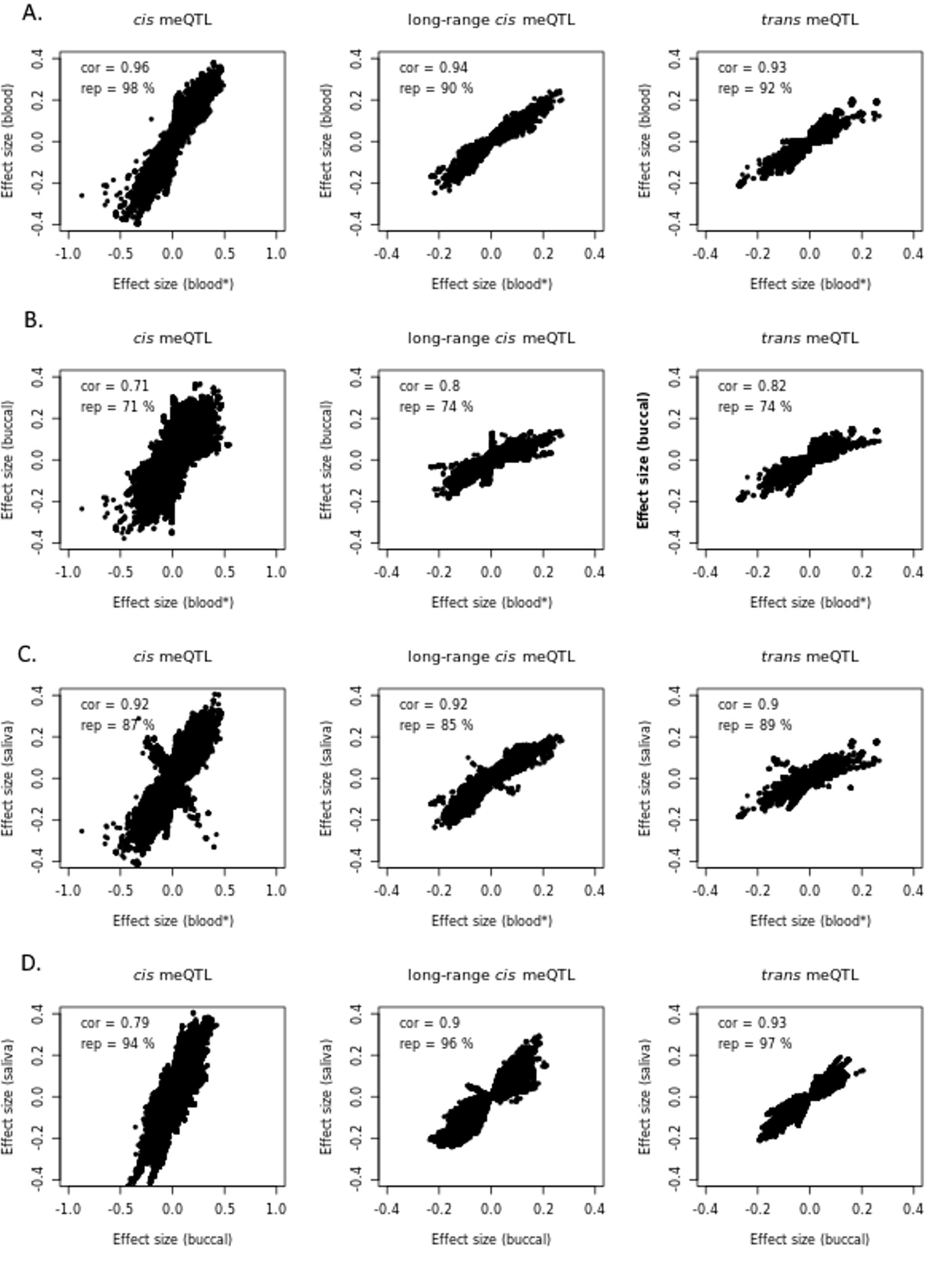
Correspondence of effect sizes from SNP-CpG pairs identified by meQTL analysis. A-C: Results from Hawe et al.^22^ (blood*) compared to blood from this study (A), buccal (B) and saliva (C). D: Effect sizes of meQTL analysis in buccal tissue compared to saliva specimens using data generated in this study.

Next, we calculated how many of the genome-wide significant SNPs-CpGs pairs correspond across all three and at least two out of three tissue types and what proportion of SNPs-CpGs pairs is present in only one tissue (see Supplementary Figure S3). We identified a much larger proportion of *cis* meQTLs (67%) that are not tissue-specific vs. those that are tissue-specific (6%). This observation is similar to published cross-tissue meQTLs results^29^, although these did not investigate buccal or saliva samples. Overall, 67/78/86% of the *cis*/lr-*cis*/*trans* SNP-CpG pairs, respectively, showed significant signals in all three tissues. This suggests that meQTL associations become less tissue-specific with increasing distance between CpG site and SNP.

### Identification of long-range cis and trans meQTL regions shared across tissues

Previous work suggested that blood meQTL SNPs acting in *trans* often regulate a large number (several hundreds to thousands) of CpGs located in the same functional unit (i.e. gene) of the genome arguing for shared molecular effects^22^. With the data from our study we were able to independently assess these findings in blood and extend them to buccal and saliva specimens. To this end, we first annotated *trans* meQTL SNPs to genes (Methods), which resulted in 9,302 genes in blood (vs. 6,816 in buccals, 7,154 in saliva). We then examined whether the 20 most frequently annotated genes were also present in the 162 (∼top 2% from 9302 genes) most frequently annotated genes in the other tissue types (see **Table 2** and Supplementary Table S4 and S5). In general, all “top-20” genes from one tissue are observed to occur among the top ∼2% (n=162) genes of the other tissues, except *NFKB1*, which had no significant *trans* meQTL in buccal tissue, and *RP11-876N24.2* which did not show up in blood. Conversely, *trans* meQTL SNPs were most frequently annotated to *MAD1L1* in all three tissues.

**Table 2.**
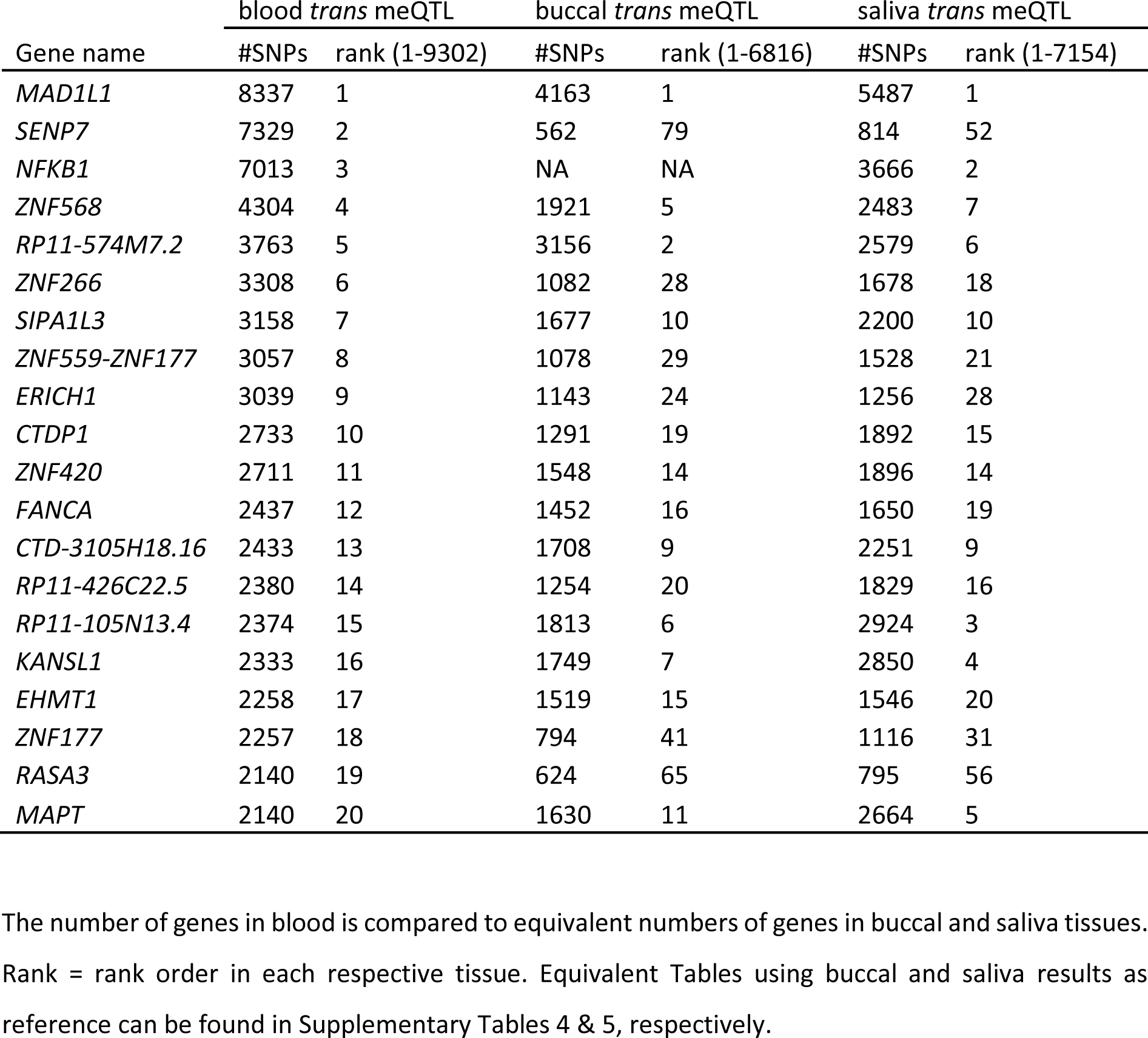
The 20 genes most frequently annotated to genome-wide significant SNPs representing *trans* meQTL in blood.

Analogously, we examined genes acting in long-range *cis* regions. For these SNPs, we identified 3610 genes in blood (vs. 3189 in buccal, 3162 in saliva). Similar to *trans* regions, all top 20 genes range among the top signals in all three tissue types, i.e. all are observed in the top 2% (often even among the top 50) genes of the other tissues (Supplementary Tables S6-S8). The meQTL SNPs in the lr-*cis* region were most frequently annotated to *MSRA* and *RP11-574M7.2* in all three tissue types.

Additionally, and following a similar visualization as in Hawe et al. ^22^, we annotated the top 1% of all significant *trans* SNPs to the most frequent meQTL genes within the +/-10 Mb region, to identify top independent regions (**Figure 4A-C)**. This annotation revealed blood to have the highest number of independent regions (n=22), followed by buccal tissue (n=21) and saliva (n=16). While many of the meQTL act in a cell type dependent manner, i.e. a substantial proportion of the annotated top 1% independent regions in blood is not included among the independent top 5% regions in buccals and saliva (i.e. 10/22= 45% and 2/22=9%, respectively), the results of the blood meQTL analyses are highly similar to those from Hawe at al. ^22^. In that study, 12 top independent regions were annotated, of which only one (*LINC00273*) is not included in the top 5% independent regions from our blood meQTL GWAS. A similarly high correspondence rate was observed within the two independent buccal datasets, where all top 1% regions in BASE-II are included in the top 5% of independent regions from the BBHI-LCBC-buccal dataset (Supplementary Figures S2).

**Figure 4.**
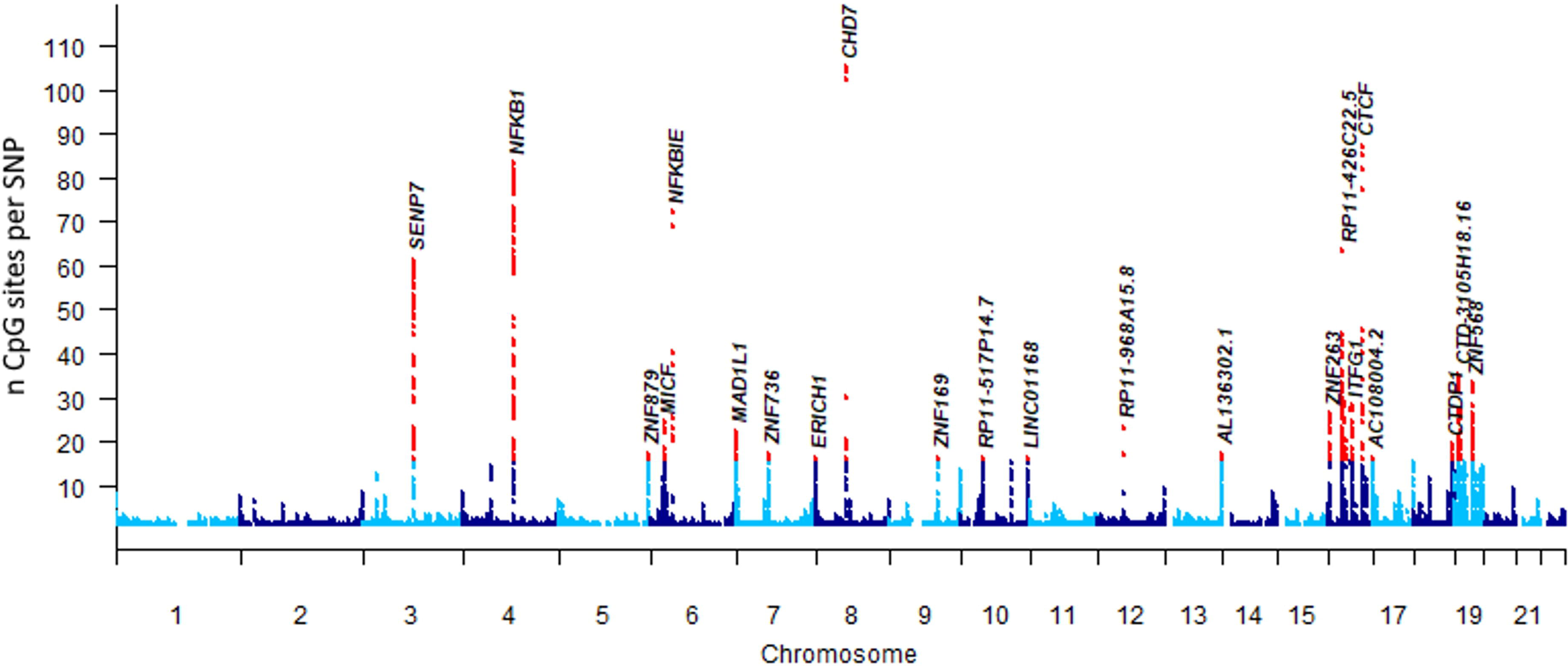
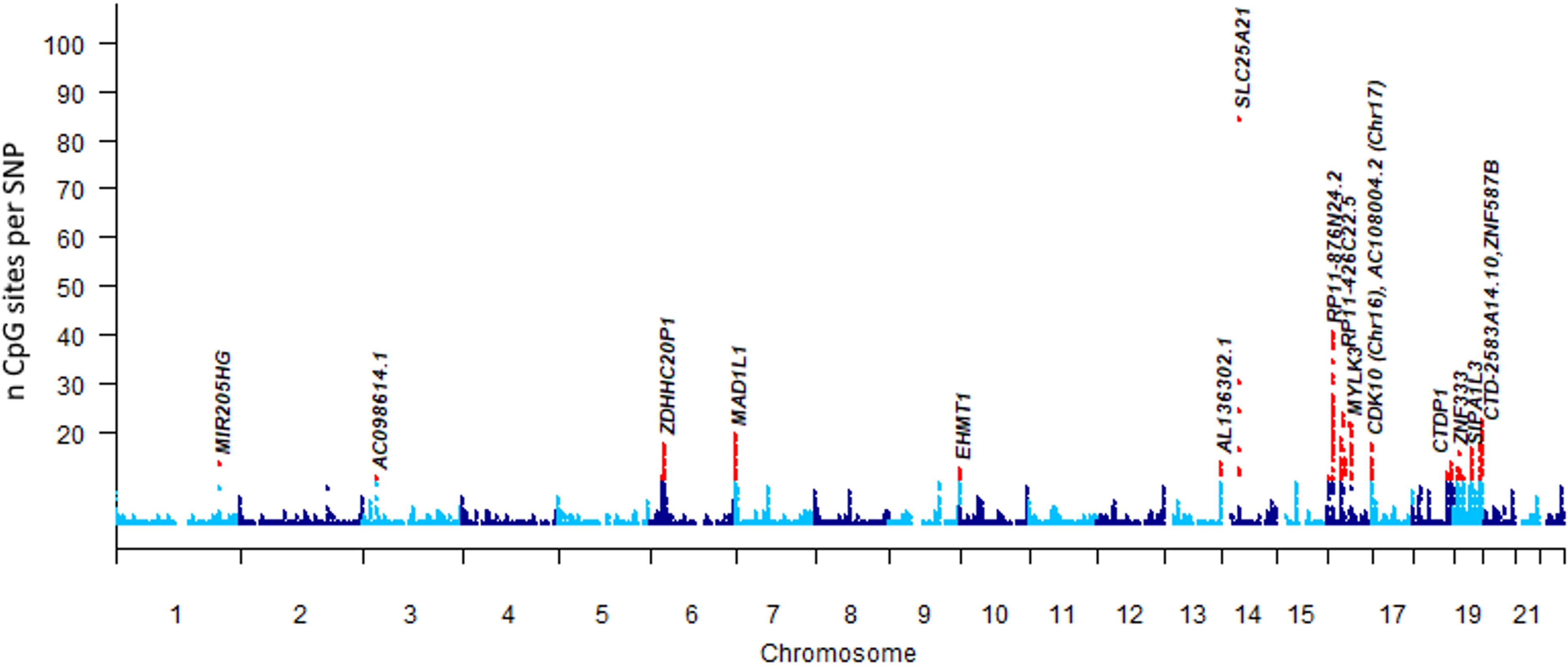
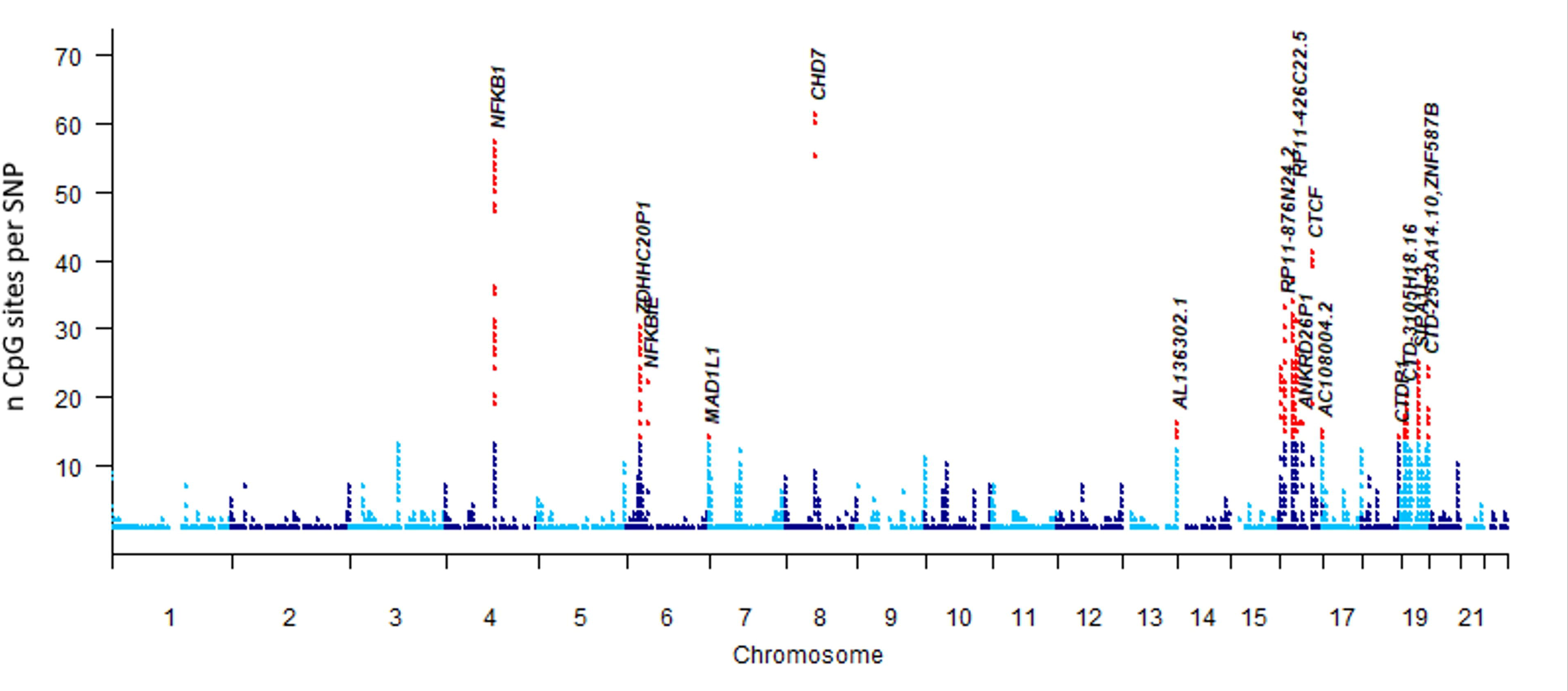
Manhattan plots of *trans* acting SNP-CpG associations in A. blood (i.e. BASE-II; n=1058), B. buccal mucosa (i.e. BASE-II, LCBC and BBHI; n=1527), and C. saliva (i.e. LCBC; n=837) dataset. Each dot represents a SNP marker. Genomic location of SNPs is on the X-axis, the number of CpG sites associated in *trans* with each SNP are on the Y-axis. Red dots indicate the top 1% SNPs across all detected trans associations. Gene names are provided for regions of the top 1% SNPs. SNPs are annotated to most frequent meQTL genes in +/-10 Mb region and in 1% all top SNPs. When two genes are equally frequent, we chose the gene previously reported in Hawe at al.^22^, when present.

### Causal relationships between DNA methylation and neurodegenerative diseases using summary data-based Mendelian randomization (SMR) analysis

In the next stage of our project, we addressed the question whether there is a causal link between DNAm and AD/PD risk using *cis* meQTL SNPs as instrumental variables via MR analyses. Disease-specific risk effects and association evidence was extracted from two recent GWAS on the respective disorders, i.e. the study by Bellenguez et al. (n=1,474,097)^1^ for AD and by Nalls et al. (n=788,989)^2^ for PD. We used the SMR tool to prioritize significant SNP-CpG signals identified in our meQTL GWAS analyses for follow-up using two-sample MR analyses (next section).

Overall, 118,757 SNPs overlapped in the AD GWAS and blood meQTL GWAS summary statistics and could be used in SMR (total n=311,882 unique genome-wide significant meQTL CpGs in *cis* across all tissue types). In blood, SMR results suggest a potential and study-wide (using a Bonferroni-corrected α of 0.05/311,882=1.6×10^-7^) significant causal relationship between DNAm and AD at 220 SNP-CpG pairs (Supplementary Tables S9). Of these pairs, 64 (of 220) show evidence for a single shared underlying causal variant affecting both DNAm in blood and AD risk (i.e. P>0.05 in the HEIDI test). These 64 SNP-CpG pairs map to 42 independent loci (25 map to genes, while 17 are located in intergenic regions; Supplementary Table S9). Equivalent analyses for the meQTL analyses in buccal (saliva) tissue prioritized 156 (176) study-wide significant SNP-CpG pairs and 33 (12) show evidence for a single shared underlying causal variant across 24 (11) loci (Supplementary Tables S10-S11; Supplementary Figure S4).

For PD, a total of 118,945 SNPs overlapped between meQTL in blood and the PD risk GWAS and were used in the SMR analyses. In blood, we identified 114 SNP-CpG pairs with a potential causal relationship with PD (p<1.6×10^-7^; Supplementary Tables S12-S14) and 13 SNP-CpG pairs in blood showed evidence of a single shared underlying causal variant affecting both DNAm and PD risk (i.e. P>0.05 in the HEIDI test). These 13 SNP-CpG pairs map to 13 loci (6 map to genes, while 7 are located in intergenic regions; Supplementary Table S12). Of all 114, equivalent SMR analyses for the PD and meQTL analyses in buccals (saliva) prioritized 101 (101) study-wide significant SNP-CpG pairs of which 10 (7) show evidence for a single shared underlying causal variant across 10 (7) loci (Supplementary Tables S12-S14; Supplementary Figure S5).

### Follow-up of SMR results by two-sample Mendelian randomization (MR) analyses

Genes identified as significant by SMR are not necessarily causally related to the phenotype in question, they merely stand a higher chance of being in such a relationship (hence our use of SMR as a “prioritization” approach)^30^. Potential causal relationships were assessed by two-sample MR using the genes / loci prioritized by SMR using the MendelianRandomization tool^31^. In addition, we tested the top 10 CpGs emerging from the meQTL GWAS analyses for *cis*, lr-*cis* and *trans* loci in each of the three tissues (i.e. an additional 10×3×3=90 CpGs) as these capture particularly strong genetic effects on DNAm at these sites that may be missed by the standard SMR paradigm. A summary of our MR workflow and numbers can be found in Supplementary Figure S6.

For AD, we identified 42 independent regions in blood (24 in buccal and 11 in saliva; p<1.59×10^-7^ & pHeidi>0.05) using SMR analysis. For MR analysis, we utilized a larger CpG selection to include all CpGs in the “prioritized” region, based on inclusion of all CpGs mapped to each region for which there is at least one significant meQTL SNP (p<10^-14^). This selection procedure resulted in 297 prioritized CpGs in blood tissue (214 in buccal, 60 in saliva). For each of these CpGs, we performed two-sample MR analysis if at least 3 independent meQTL SNPs (r^2^<0.1, 1000 kb and P<10^-14^) overlapping with SNPs from the respective GWAS summary statistics and not identified as outliers using the MR-PRESSO tool^32^ were available. Overall, this procedure resulted in sufficient data for a total of 193 MR analyses in AD (Supplementary Figure S6). In total, we identified nine (four in blood and five in buccal tissue) putative causal CpGs with p-values falling below the multiple-testing corrected threshold for this part of our study (p<1.47×10^-4^; Supplementary Figure S6) in all four MR models tested (Supplementary Figure S7). One example is *KANSL1*, which showed highly significant evidence for a causal relationship (i.e. a positive sign in the effect size estimate) with AD risk in blood (cg09860564, smallest p = 4.08×10^-13^) and buccal tissue (cg17642057, smallest p = 1.21×10^-12^). *KANSL1* is functionally interesting as it maps to an inversed haplotype region on chr. 17q21 into the immediate vicinity of the gene encoding microtubule associated tau protein (*MAPT*), whose accumulation as neurofibrillary tangles represents a neuropathological hallmark of AD^1^. The other significant two-sample MR signals were elicited by individual CpG sites in *PSMC3* (blood; smallest p = 1.2×10^-12^) and *PRDM7* (buccal; smallest p = 9.6×10^-13^), as well as two CpGs in *TSPAN14* (buccal; smallest p = 2.72×10^-21^). In addition, there were three (of which two were observed in blood) CpGs from intergenic regions showing significant effects (Supplementary Figure S7).

In PD, the equivalent number of CpGs assessed by two-sample MR analyses was 146 (Supplementary Figure S6). From these, we identified 15 putative causal CpGs showing study-wide significant evidence for potential causal effects of DNAm on PD risk across all three analyzed tissues (Supplementary Figure S8). Interestingly, all PD CpGs were located in the inversed haplotype region on chr. 17q21 in and near *MAPT* (within a ∼500kb window encompassing *CRHR1*, MAPT, and *KANSL1*). Of note, none of the significant 17q21 CpGs in PD overlapped with those that emerged in this region for AD. No other PD loci outside the 17q21 region were highlighted by two-sample MR using our novel meQTL catalogs.

### Sensitivity analyses on the two-sample Mendelian randomization results

MR results can be biased towards false-positive findings by residual correlation between SNPs used as “independent” instrumental variables^33^. The default correlation (i.e. linkage disequilibrium) threshold used in the primary analyses here was r^2^≤0.1, which represents a commonly applied cut-off in this type of MR setting^19^. To assess the stability of our MR results and to minimize potential bias due to residual correlation among SNPs, we recalculated all significant two-sample MR results using more stringent correlation thresholds, down to r^2^≤0.01 (Methods). As can be seen from Supplementary Tables S15 & S16, the number of usable independent SNPs dropped below the recommended minimum of three for many CpGs and only left one CpG each (AD: cg20307385 in *PSMC3* [**Figure 5**; Supplementary Table S15]; PD: cg07936825 in *MAPT* [Supplementary Table S16]) for the MR analyses at the most stringent threshold of r^2^≤0.01. Both of these CpGs continued to show strong and consistent association by MR across all four models used. Using additional r^2^ thresholds between 0.1 and 0.01 enabled additional sensitivity MR analyses for a total of 13 out of 28 CpGs (Supplementary Tables S15 & S16). In the majority of cases, the initial MR results were confirmed, although many showed less significant effects, likely owing to the lower number of instrumental variables (i.e. SNPs) available at these more stringent thresholds. Only the MR results for four CpGs (1 AD, 3 PD) yielded non-significant (p>0.05) results in at least one MR model using an r^2^ threshold <0.1 (Supplementary Tables S15 & S16). While this could indicate a possible bias in the primary MR analyses at these CpGs, we emphasize that non-significance only affected the “simple model” in each instance and the support remained highly significant even for these four CpGs in the three remaining MR models. Thus, by and large, these sensitivity analyses do not indicate the presence of a strong bias in our MR results, at least not for the CpGs where additional testing was possible.

**Figure 5.**
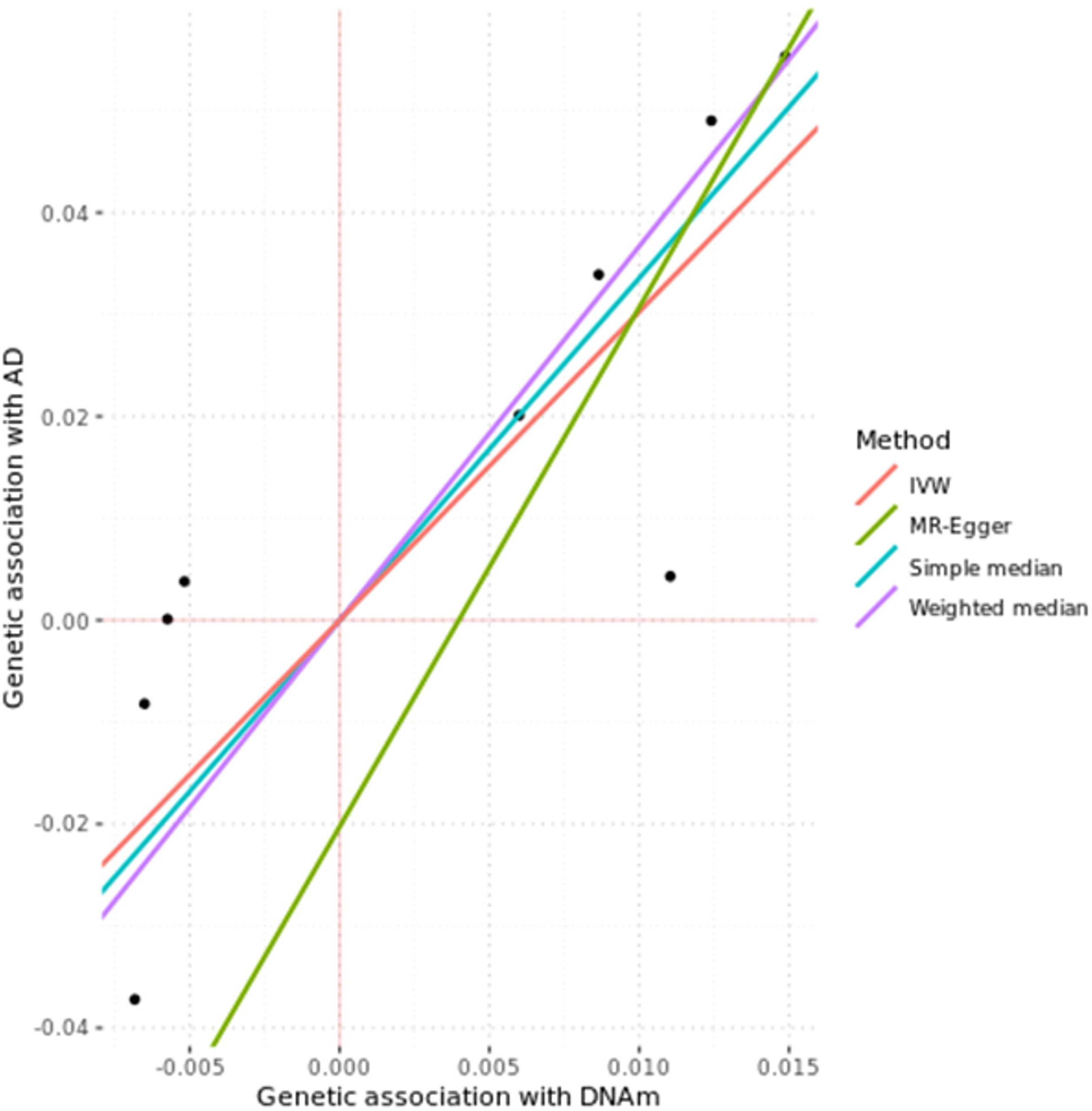
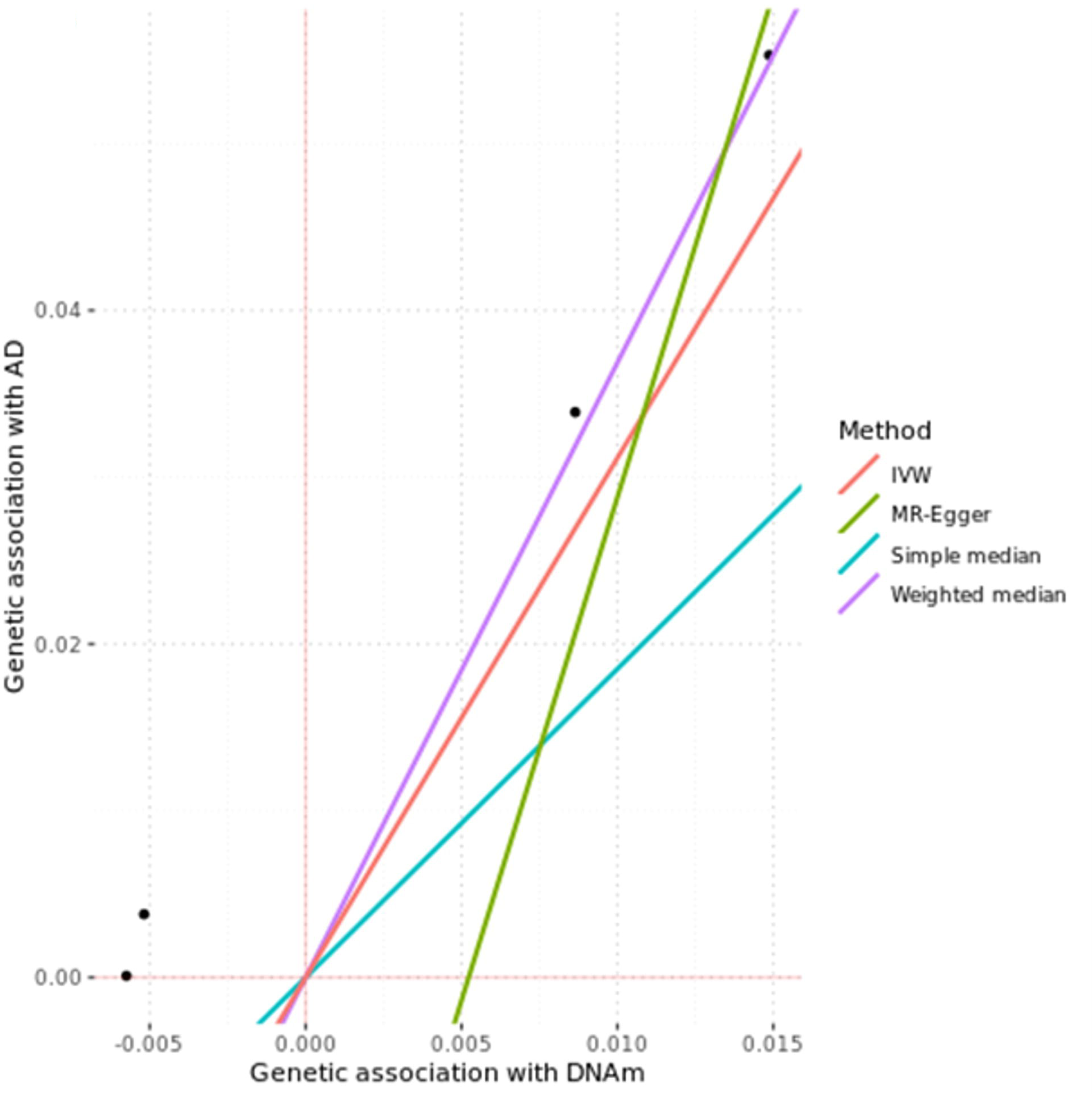
Example MR analysis using four models estimated causal link to cg20307385 in *PSMC3* in AD using independent instrumental variables (SNPs) at A. r^2^<0.1., and B. r^2^<0.01. Effect sizes and p-values corresponding to these analyses are depicted in Supplementary Table S15.

As an additional line of sensitivity analyses, we tested for colocalization of GWAS results, i.e. SNPs representing both meQTLs and risk variants. In general, evidence for colocalization (which indicates that the same variant is driving the meQTL and disease associations) is regarded as supportive of a significant MR finding^19^. In AD, support for colocalization was observed for four of the nine MR CpGs (Supplementary Table S17). This relates to CpGs in *PSMC3* (1 CpG) and *TSPAN14* (2 CpGs) as well as an intergenic probe on chromosome 11q14 (cg04441687), which is located approx. 100kb downstream of *PICALM*. Thus, for these four CpGs, all statistical evidence accrued in the multipronged analyses performed in this study unequivocally points to a causal relationship between DNAm and risk for disease. Interestingly, all four of the implied regions also show evidence for differential methylation in AD vs. control brain samples in the recent brain EWAS meta-analysis by Smith et al.^11^ (Supplementary Table S17 and next section). In PD, where we identified a large number of consistent and highly significant two-sample MR signals for CpG sites in a ∼400kb region on chromosome 17q21 (encompassing *CRHR1-MAPT-KANSL1*), none of the colocalization results favored the presence of a shared variant (H4), but in all instances pointed to the existence of two separate variants underlying DNAm and PD risk (H3; Supplementary Table S18). The most obvious scenario in which such “conflicting” results can occur is that the exposure and outcome have distinct causal variants that are in linkage disequilibrium, which in return may signify that one of the MR assumptions may be violated^19^.

### Comparison of novel meQTL-based MR results and brain-based EWAS for AD and PD

As outlined in the introduction, the application of MR to imply causal relationships between exposure (here: DNAm) and outcome (here: AD/PD risk) can effectively circumvent the problem of “causality uncertainty” of EWAS. In this regard our study leverages the oftentimes substantially larger sample sizes typically used for disease risk GWAS (here ranging from 788,989 in PD^2^ to 1,474,097 in AD^1^) and meQTL GWAS (here exceeding 1,500 samples for blood). The sample sizes for the largest (to our knowledge) primary EWAS in brain samples for AD are n∼960^11^ and n∼320 for PD^17^. Notwithstanding the (much) reduced power of these primary EWAS, evidence for differential DNAm at overlapping loci from these studies may still be regarded as supportive of the MR-based findings derived from our data. To this end, it is comforting to note that four of the eight top MR-based loci identified here are also supported by EWAS, at least in AD (Supplementary Tables S17 [AD] and S18 [PD]).

First, *TSPAN14* was reported as one of the top EWAS findings in the recent meta-analyses performed by Smith et al.^11^. In that study, the authors found evidence for differential DNAm at cg16988611 (p=1.9×10^-10^ in prefrontal cortex, and p=9.98×10^-12^ in the cross-cortex analyses). While the two lead CpGs in this locus in our study (i.e. cg24699150 and cg22345419) were not analyzed by Smith et al.^11^ since neither probe is included on the 450K array, it is reasonable to assume that these two signals represent the same underlying effect. Second, while for *PSMC3*, Smith et al.^11^ reported no significant results for our primary CpG (cg20307385), they did find genome-wide significant evidence for association with cg06784824 (p=3.0×10^-8^) which is located ∼75kb proximal in the last exon of *SPI1*. In the most recent AD risk GWAS^1^, this general locus is annotated to extend from *SPI1* to *CELF1*, and *PSMC3* is one of five genes mapping into this interval (see Supplementary Fig. 16 in ref. ^1^). This could indicate that there are two independent DNAm sites acting in this region (one near the 3’ end of *SPI1* and one within *PSMC3*) or that they are pointing to the same signal that is also highlighted by the AD GWAS. Third, our MR signal near *PRDM7* (elicited by cg16611967) is directly confirmed by Smith et al.^11^, who report at least nominal (p<0.05) evidence for differential methylation with this probe in their cross-cortex EWAS analyses. The much stronger statistical support here (p-values ranging from 3×10^-3^ to 6×10^-13^; Supplementary Table S15) is likely afforded by the much larger sample sizes, and hence power, of our analyses. Fourth, our MR signal on chromosome 11q14 elicited by cg04441687 ∼200kb upstream of *PICALM* cannot be directly compared with Smith et al.^11^ since this CpG site is missing from the 450K array. However, they report cg07180834, which maps approx. 30kb p-ter from our signal, to be differentially methylated in prefrontal cortex (p=0.001), implying the same general region surrounding our finding. Lastly, Smith et al.^11^ report genome-wide significant evidence for differential methylation with at least two CpGs in the general *MAPT* region on chromosome 17q21, i.e. cg20864568 (p=9.93×10^-8^ in prefrontal cortex) and cg15194531 (p=1.74×10^-8^ in the cross-cortex analyses), suggesting that the link between this locus and AD risk may, indeed, be mediated by differences in DNAm. While our MR results for this locus clearly imply causality underlying this link, we note that the missing evidence for colocalization (see previous section) may indicate some bias in the MR analyses.

While for PD essentially all of our MR results could be directly compared to the EWAS by Pihlström et al. ^17^ who also used the EPIC array, none of our PD-related results within the *KANSL1/MAPT* region on chromosome 17q21 showed evidence for differential DNAm in that study. This may, at least partially, be due to the exceedingly small sample size (n∼320) used in that EWAS.

In summary, there is either direct or indirect support for five of our seven MR-based DNAm loci from a recent AD EWAS using samples from different human brain regions. This can be seen as evidence for independent validation on two levels: i) Validating the overall approach taken here, i.e. combining peripheral (non-brain) meQTL data with AD genetics to derive mechanistically relevant links acting in the brain, and ii) for the involvement of DNAm underlying the well-established AD risk associations at these five loci. We note that the non-confirmation by EWAS of the remaining two intergenic CpGs (cg04043334 at 10q23 and cg02521229 at 11q12) does not necessarily imply the absence of a causal relationship owing to the much smaller genomics resolution of the primary EWAS^11^. We also note that the former of these two intergenic CpGs (cg04043334) maps ∼143kbp of *TSPAN14*, which is one of the major brain EWAS signals in the study by Smith et al.^11^

## Discussion

In this work, we performed extensive genome-wide mapping of meQTLs in three peripheral human tissues of which two (buccal and saliva) were not sufficiently covered in comparable previous efforts. After performing more than 12 trillion statistical tests, we identified between 11 and 15 million genome-wide significant SNP-CpG pairs in each tissue. Most of these (∼90%) were located in *cis* while long-range *cis* and *trans* effects comprised approx. 5% each of the remaining signals. In a second step, we combined these novel meQTL GWAS results with large risk GWAS for AD and PD using a multipronged MR and colocalization analysis approach to assess whether any of the hitherto reported AD/PD GWAS signals might unfold their effects by affecting DNAm. These analyses strongly suggest that the GWAS-based risk associations between *PSMC3*, *PICALM*, *TSPAN14* in AD may (at least partially) be due to differential DNAm at or in the vicinity of these genes. In addition, there is strong – albeit less unequivocal – support for causal links between differential DNAm at *PRDM7* in AD as well as at *KANSL1/MAPT* in AD and PD. To facilitate like-minded analyses in other complex human traits, we made the complete and entirely novel meQTL GWAS results freely available to the scientific community (URL: https://doi.org/10.5281/zenodo.10410506).

Our study has several strengths, which include: i) utilizing comparatively large sample sizes across the three different tissue types (resulting in the largest meQTL GWAS ever performed in buccal and saliva specimens); ii) using the highest resolution DNAm microarray currently commercially available; iii) employing stringent statistical thresholds to declare genome-wide significance; iv) assessing and establishing independent replication for top meQTLs in buccal and blood tissue; and v) applying a multipronged and state-of-the-art analysis approach to infer potential causality between DNAm and disease associations for two common neurodegenerative disorders each based on the largest risk GWAS published in the respective fields.

Despite these strengths, there are a number of caveats and potential limitations inherent in our study. First, all identified associations, including meQTL results and potentially causal disease links, are of a statistical nature and do not necessarily imply true molecular relationships. While we went to great lengths to limit false-positive or biased results throughout the various analysis arms of our work, none of the reported associations should be regarded as established until further validation from functional experiments. We note, however, that at least for buccal and blood tissue we observe very high (>>90%) replication rates for our top meQTL findings suggesting that the genetic effects on DNAm in these tissues are relatively stable and likely genuine. Second, by design, the meQTL maps provided are limited by the resolution of the utilized DNAm microarray. While this covers ∼850K CpGs in functionally relevant regions of the human genome, this number still only represents ∼1/30^th^ of the CpG sites that can be measured by whole-genome DNAm sequencing approaches. While this difference in resolution is substantial, the use of high-throughput microarrays is much more cost efficient allowing to assay much larger numbers of samples and, hence, to achieve greater statistical power. Third, performing and interpreting MR analyses for causal inferences has many drawbacks and potential limitations (extensively discussed in refs. 19,20). Again, we approached this topic with great caution by performing a large number of alternative and supporting analyses to derive the best possible inferences in the context of our study. However, we cannot exclude the possibility that some (or even all) of our MR-based conclusions may be biased or false. Only dedicated molecular experiments directly testing the hypotheses put forward in our study can help to distinguish true from false causal links. Finally, both the newly derived primary meQTL maps as well as the AD/PD risk GWAS are based on individuals of European descent. Therefore, no inferences can be made with respect to potential causal links highlighted here in individuals of different ethnicities.

In summary, our study represents a tour de force analysis resulting in the largest and most comprehensive catalogue of meQTL effects in human buccal and saliva tissues. Using these and additional blood-based meQTL data implicates a likely causal role of differential DNAm in AD and PD development in some genomic regions associated with disease risk by GWAS. Future work needs to independently replicate our results and elucidate the molecular mechanisms underlying these associations.

## Methods

### 1. Methylation quantitate trait locus (meQTL) genome-wide association study (GWAS) and independent replication analyses

#### 1.1 Human samples

In this study, we analyzed a total of 2,592 samples from three independent datasets (“Berlin Aging Study II” [BASE-II] recruited in Berlin, Germany, “Barcelona Brain Health Initiative” [BBHI] recruited in Barcelona, Spain, and “Lifespan Changes in Brain and Cognition” [LCBC] recruited in Oslo, Norway) collected under the auspices of the EU-funded Lifebrain study^34^. Lifebrain participants for this study were selected based on the parallel availability of genome-wide SNP genotype and genome-wide DNA methylation data. Supplementary Table S19 provides a summary of demographic characteristics of the datasets used in this study. The use of DNA samples for genomics and epigenomics analyses in Lifebrain was approved by the ethics committee of University of Lübeck (approval number: 19-391A).

##### Berlin Aging Study II (BASE-II)

The portion of the BASE-II dataset used in this study consists of adult residents (age range: 23-88 years) from the greater metropolitan area of Berlin, Germany ^35,36^. For this study, we included DNA samples collected from blood (n=1,058) and buccal swabs (n=837) collected at the second examination conducted between 2019-2020 as part of the “GendAge” project^36^. Of these, a total of n=830 BASE-II participants contributed DNAm data from *both* blood and buccal swabs, and this overlap was taken into account in analyses assessing the correspondence of meQTL effects in these two tissues (Supplementary Table S19). The BASE-II/GendAge studies were conducted in accordance with the Declaration of Helsinki and approved by the ethics committee of the Charité— Universitätsmedizin Berlin (approval numbers: EA2/144/16, EA2/029/09).

##### Barcelona Brain Health Initiative (BBHI)

The Barcelona Brain Health Initiative (BBHI) is an ongoing, longitudinal study recruiting participants from the greater metropolitan area of Barcelona, Spain, with the focus on evaluating factors determining brain health^37^. For this study, there were buccal samples from n=372 BBHI participants (age range: 30 to 67 years) available which were subjected to genome-wide SNP and DNAm profiling (Supplementary Table S19). Collection of BBHI samples was conducted in accordance with the Declaration of Helsinki and following the recommendations of the “Unió Catalana d’Hospitals” with written informed consent from all subjects. The protocol was approved by the Unió Catalana d’Hospitals (approval number: CEIC 17/06).

##### Lifespan Changes in Brain and Cognition (LCBC)

This dataset comprises a collection of n=1,155 participants (age range: 20-81) recruited mostly in the larger metropolitan area of Oslo, Norway, by investigators at LCBC, as well as through other collaborations within Norway. The sample comprises phenotypically well-screened individuals with comprehensive neuropsychology, MRI, lifestyle, health, biomarkers, and other measures. For this study, DNA extracts used for genome-wide SNP genotyping and DNAm profiling originated from n=318 buccal swabs and n=837 saliva samples, which were non-overlapping (Supplementary Table S19). The studies were approved by the Regional Ethical Committee of South East Norway. Written informed consent was obtained from all participants.

#### 1.2 Genome-wide SNP genotyping, quality control, and imputation

DNA for all samples was extracted using standard procedures as described previously (BASE-II & BBHI: ref. 14 ; LCBC: ref. 38). Genome-wide SNP genotyping was performed using the Global Screening Array (GSA; Illumina, Inc., USA) at the Institute of Clinical Molecular Biology at UKSH Campus Kiel on an iScan instrument according to the manufacturer’s recommendations. Genotype calling, quality control (QC) and imputation steps were performed using an automated bioinformatics workflow described previously^39,40^. Briefly, genotype determination from the raw intensity data was performed using GenomeStudio (Illumina, Inc., version 2.0.2), QC with the PLINK program (version 1.9) ^41,42^. Imputation of untyped variants was performed with MiniMac3 ^43^ software using the “Haplotype Reference Consortium” (HRC; v1.1 [EGAD00001002729 including 39,131,578 SNPs from ∼11K individuals]) reference panel ^44^. Finally, allele dosages (i.e. genotype probabilities) of ∼39 million SNPs per proband were available for post-imputation QC. This entailed filtering at both the SNP and individual levels using the following criteria. <u>SNP-filtering</u>: SNPs were excluded with low imputation quality score (r^2^ < 0.7), minor allele frequency (MAF) below 5%, genotyping rate below 98%, and significant deviations from Hardy-Weinberg Equilibrium (HWE) in control individuals (p < 5 × 10^-6^). <u>DNA sample filtering</u>: individual-level genotyping data were excluded in case of low genotyping efficiency (< 98%), discrepancies between genetic and clinical recorded sex, duplicated DNA samples, cryptic relatedness (--king-cutoff 0.025), and samples with implausible heterozygosity (mean ± 6 × SD). To correct for population stratification, genetic ancestry was mapped onto genotype data from the 1000 Genomes project (using that study’s five “superpopulation” codes), followed by principal component analysis (PCA) performed in PLINK (v2.0). Only samples clustering to the “CEU” population cluster were retained for analyses. Genomic location of SNPs throughout this manuscript are based on human genome build GRCh37/hg19.

#### 1.3 DNA methylation measures and quality control in blood, buccal and saliva tissues

Genome-wide DNA methylation (DNAm) profiles were generated in the same individuals whose samples were also used in the genotyping experiments. DNAm was measured using the Infinium Human MethylationEPIC array (Illumina, Inc., USA) at the Institute of Clinical Molecular Biology at UKSH Campus Kiel on an iScan instrument according to the manufacturer’s recommendations. QC and data processing were performed using the same procedures as described previously ^12,14^. Briefly, data pre-processing was performed in R (version 3.6.1) using the package *bigmelon* (version 1.22.0) with default settings^45^. Cell-type composition estimates were obtained with the R package EpiDISH (version 2.12.0)^46^ and used for correction of DNAm β-values. Samples were excluded from the analysis if (a) the bisulfite conversion efficiency was below 80% according to the bscon function in the bigmelon package, (b) the sample had a beadcount < 3 in more than 5% of all probes, (c) the sample had a detection p-value below 0.05 in more than 1% of all probes, (d) the sample was identified as an outlier according to the outlyx function in the bigmelon package using a threshold of 0.15, (e) the sample showed a large change in β-values after normalization according to the qual function in the bigmelon package with a threshold of 0.1, (f) the sample showed a discrepancy between predicted sex according to the Horvath multi-tissue epigenetic age predictor and reported sex, or (g) there was a greater than 70% discrepancy between genotypes of 42 SNPs determined concurrently from the EPIC and GSA SNP genotyping array. All samples were normalized with the dasen function of bigmelon. Further details on QC and data processing can be found in refs. 12 and 14 from our group. Genomic location of CpGs throughout this manuscript are based on human genome build GRCh37/hg19.

#### 1.4 Identification of genetic factors influencing methylation, (meQTL GWAS)

After QC of both genome-wide SNP genotypes and DNAm patterns, we performed meQTL genome-wide association analyses separately for blood, buccal mucosa, and saliva using the R package MatrixeQTL^47^. In detail, we applied linear regression models including sex, the first ten principal components of a PCA assessing genetic ancestry, and the first five principal components of DNAm levels at pruned CpGs as covariates (see ref. ^12,48^ and supplementary methods). For the buccal datasets an additional dummy variable was introduced to adjust for laboratory batches. We retained only test statistics from SNP-GpG pairs showing p-values less than 0.05. To account for multiple testing, we applied the same study-wide threshold as in Hawe et al.^22^ (i.e. α = 1×10^-14^), which was based on the ∼4.3 trillion tests performed in that study conducting a meQTL GWAS in one tissue (blood). While here we performed approximately three times the number of tests owing to the analysis of two additional tissue types, we note that not all of these were independent owing to the correlation structures between SNPs (in Europeans there are approximately 1M independent SNPs at MAF≥0.05) and CpGs (the EPIC array contains approx. 530K independent CpGs^49^. Assuming that the meQTL effects are completely independent across the three tissues used in our study, this would amount to a total of 3×1Mx530K=1.7×10^12^ independent tests and an effective Bonferroni-corrected α-level of 0.05/1.59×10^12^ = 3.1×10^-14^ which is slightly less conservative than the level actually applied (i.e. α = 1×10^-14^). Following the analyses by Hawe et al.^22^, we separated our findings into *cis* meQTL (SNP-CpG distance within 1 Mb), lr-*cis* meQTL (>1 Mb apart but on the same chromosome) and *trans* meQTL (associations between SNPs and CpG sites on different chromosomes).

#### 1.5 Independent replication analyses

To assess the replicability of meQTL results in buccal tissue, we were able to use two independent datasets in this study: one discovery dataset containing samples from the Berlin Aging Study II (BASE-II) and another replication dataset containing samples from the Barcelona Brain Health Initiative (BBHI) and the Centre for Lifespan Changes in Brain and Cognition (LCBC). For the meQTL GWAS, BBHI and LCBC were analyzed jointly adjusting for center using a dummy variable. We assumed that a meQTL from the discovery dataset was replicated if it showed evidence of association at P values <0.05 and consistent direction of effect in the replication dataset.

For meQTL results replication in blood tissue, we have downloaded the results from Hawe et al.^22^ who performed a meQTL GWAS in blood identifying of 11,165,559 study-wide significant meQTLs. To calculate the replication rate, we considered only the SNPs and CpGs that also remained in our analysis after QC. For the SNP-CpG pairs with a P value above 0.05, the test statistic was recalculated and saved. We considered a meQTL as replicated if they showed evidence for association at p-values <0.05 and an effect direction consistent with that reported in Hawe et al.^22^.

For salivary tissue, we could not estimate the replication rate because no additional data were available.

### 2. Comparison of meQTL findings across three tissue types: blood, buccal, saliva

For comparison, we used meQTLs results from Hawe et al ^22^ to estimate the replication rate and correlation of effect estimates between the different tissues. These previous results were compared to post-QC SNPs-CpGs pairs from each dataset analyzed here (i.e. buccal and saliva). For the SNP-CpG pairs with a P value above 0.05, the test statistic was recalculated and saved. We considered a meQTL as replicated if they showed evidence for association at p-values <0.05 and an effect direction consistent with that reported in Hawe et al.^22^. Furthermore, we calculated the correlation of meQTL effect estimators using Pearson’s method.

### 3. Identification of long-range cis and trans meQTL regions shared across tissues

We identified the shared regions by annotating the significant meQTL SNPs to genes using ANNOVAR software^50^ based on their physical position on the chromosomes (hg19/GRCh37) as provided on the UCSC genome browser (http://hgdownload.cse.ucsc.edu/goldenPath/hg19/database/ensGene.txt.gz). For comparison of tissue-specific regions at the SNP level, we estimated the top 1% SNPs across all detected long-range *cis* and *trans* associations and annotated the top SNP to the most common gene in a +/-10 Mb region. When two genes are equally frequent in one region, we chose the gene previously reported in Hawe at al.^22^, when present. We also looked at the most frequently annotated genes within all lr-*cis* and *trans* SNPs in each dataset and compared the top 20 genes within blood, buccal, and saliva tissues.

### 4. Linking DNA methylation and Alzheimer’s and Parkinson’s disease using Mendelian randomization

#### 4.1. Summary data-based Mendelian randomization (SMR) analysis

To run an initial test for association between AD/PD risk and DNAm levels across all three analyzed tissues (blood, buccal, and saliva) and to prioritize downstream two-sample MR analyses, we applied the Summary data–based Mendelian Randomization (SMR) approach^30^. Only SNP-CpG pairs attaining study-wide genome-wide significance (i.e. p-values of the top associated *cis* meQTL were <10^−14^) were considered in this analysis. The disease-specific data were retrieved from summary statistics of the two most recent GWAS meta-analyses on risk for AD^1^ (n total = 487,511) and PD^2^ (n total = 482,730). To account for multiple testing within this arm of our study, we considered the total number of unique genome-wide significant methylation CpGs in *cis*, that were included in the analysis from blood (n=118,955), buccal (n=92,694), and saliva tissues (n=100,233) yielding a total number of n= 311,882 comparisons. Accordingly, the Bonferroni adjusted α level was set to α = 0.05/311882 = 1.6×10^-7^. Whenever more than three SNPs were in linkage disequilibrium (LD; r^2^ > 0.1, 1,000 kb) with a *cis*-SNP, a heterogeneity test (HEIDI) was performed to distinguishing functional association from linkage, as implemented in the SMR-Tool^30^.

For gene prioritization from SMR, we assigned the significant CpGs to genes based on the information in the Infinium MethylationEPIC manifest file (version 1.0 B5, Illumina, Inc., USA).

#### 4.2 Systematic two-sample Mendelian randomization (MR) analyses

To test for potential causal relationships between DNAm and AD/PD, we examined SMR-prioritized regions, i.e. those showing with SMR p<1.6×10^-7^, at least one genome-wide significant meQTL SNP (p<1×10^-14^), and no evidence for significant heterogeneity (HEIDI test p>0.05) by two-sample Mendelian randomization (MR) analyses. Two-sample MR was performed using the R package MendelianRandomization ^31^ running four analysis models: simple median ^51^, weighted median ^51^, inverse variance weighted (IVW)^52^ and Egger regression ^53^. Each model makes different assumptions and uses different strategies to avoid false positive causal inferences. As recommended by the authors, we only consider those MR results further which show consistently significant signals across all four models.

In addition to testing regions prioritized by the SMR method, we also analyzed the 10 most frequently associated CpGs with SNPs in independent regions. Genes corresponding to these CpGs were annotated using the Infinium MethylationEPIC Manifest file (version 1.0 B5, Illumina, Inc., USA). CpGs in intergenic regions (i.e. not annotated to any gene) were assumed to represent one independent region each. Only independent (r^2^<0.1, 1,000 kb) SNPs present in at least one GWAS summary statistic (i.e. AD and/or PD) with a MAF >0.05 and showing p-values <1×10^-14^ in the meQTL GWAS were included in these analyses. In these high-frequency regions, MR was performed if there were at least 3 independent SNPs after outlier correction with MR-PRESSO^32^. To estimate study-wide significance for this arm of our study we used Bonferroni’s method considering the total combined number of tests performed in AD (n=193) and PD (n=146), i.e. α = 0.05/349 = 1.47×10^-4^.

#### 4.3 Sensitivity analyses

We performed an extensive sensitivity analysis using several methods. On one hand, we tested for heterogeneity, as implemented in the MendelianRandomization^31^ package and assume that instrumental variables with a p-value greater than 0.05 reject heterogeneity. On the other hand, we performed a global test to identify outliers in the data as implemented in the MR-PRESSO tool^32^. If the global test yields a p-value greater than 0.05, we assume that the data are consistent and have no local outliers. Finally, the intercept parameter is another indicator representing the average pleiotropic effect of a genetic variant^31^. If the intercept p-value remains greater than 0.05, we assume that there is no pleiotropy.

In two-sample MR analysis, the selection of correlated instrumental variables (SNPs) within a gene can lead to numerically unstable estimates of the causal effect^33^. For this reason, we recalculated the MR using squared correlation up to r^2^≤0.01 for significant CpGs identified with (r^2^<0.1, 1,000 kb) SNPs. We lowered the r^2^ threshold to the point where less than 3 SNPs remained for analysis, or we achieved an r^2^ value of 0.01.

We used colocalization as part of Mendelian randomization sensitivity analysis, testing assumptions about instrumental variables (SNPs) for a given genetic region (gene and CpGs within) using the R-package susieR^54^. In the analysis, we included all SNPs (not filtered for LD) used for MR in *cis* regions. If there is strong evidence that exposure and outcome are influenced by different causal variants, then it is implausible that variants in that region are valid instrumental variables for exposure^19^.

## Acknowledgements

The authors are grateful to all participants for their time, commitment, and willingness to participate the in BASE-II, BBHI, and LCBC studies. Part of this research was funded by the EU Horizon 2020 Grant: ‘Healthy minds 0–100 years: Optimising the use of European brain imaging cohorts (“Lifebrain”; grant #732592 to A.M.F., K.B.W., U.L., D.B.F., and L.B.), the Cure Alzheimer’s Fund (“CIRCUITS-AD” to L.B.), and the Deutsche Forschungsgemeinschaft (DFG; grant #DE842/7-1 to I.D.). Additional support was provided by the German Federal Ministry of Education and Research (for the BASE-II/GendAge studies) under grant numbers #01UW0808; #16SV5536K, #16SV5537, #16SV5538, #16SV5837, #01GL1716A and #01GL1716B. Further support came from the Norwegian Research Council (to K.B.W., A.M.F.), the National Association for Public Health’s dementia research program, Norway (to A.M.F.), the European Research Council’s Starting (grant agreement 283634 to A.M.F. and 313440 to K.B.W.) and consolidator Grant Scheme (grant #771355 to K.B.W. and #725025 to A.M.F.), the “EU Joint Programme – Neurodegenerative Disease Research 2021”(JPND2021, “EPIC4ND project” to C.M.L.) and the DFG (LI 2654/2-1 to C.M.L.). C.M.L. was supported by a Heisenberg grant of the DFG (LI 2654/4–1). Dr. D. Bartrés-Faz was partly supported by the Barcelona Brain Health Initiative and Institute Guttmann and an ICREA Academia 2019 Research Award by the Catalan Government. Dr. A. Pascual-Leone was partly supported by the Barcelona Brain Health Initiative and Institute Guttmann, the National Institutes of Health (R01AG076708, R01AG059089, R03AG072233, and P01 AG031720), and the Bright Focus Foundation. We would like to thank Dr. Johann S. Hawe at the Institute of Computational Biology, Deutsches Forschungszentrum für Gesundheit und Umwelt, Helmholtz Zentrum München, Neuherberg, Germany, for his kind assistance with generating the chessboard plots. Lastly, we acknowledge the support of the OMICS high-performance compute cluster at University of Lübeck (https://www.itsc.uni-luebeck.de/dienstleistungen/omics/omics-english.html) where essentially all computational analyses of this study were performed.

## Author contributions

1. Conception and design of study: O.O., Y.S., C.M.L., L.B.
2. Sample recruitment and handling: D.B.F., G.C., S.D., A.M.F., U.L., A.P.L., C.S.P., J.M.T., V.M.V., K.B.W., I.D.
3. Generation of molecular data: Y.S. V.D., S.S.S., T.W., M.W., A.F.
4. Statistical analysis and interpretation: O.O., Y.S., J.H., L.D., M.S., C.M.L., L.B.
5. First draft of manuscript: O.O., L.B.
6. Critical review and approval of final manuscript: All authors.

## Data availability

All results of this study have been made available on Zenodo (URL: https://doi.org/10.5281/zenodo.10410506). Together with the AD/PD summary statistics from Bellenguez et al. (ref. 1) and Nalls et al. (ref. 2) these allow a full reproduction of all MR (incl. SMR) and colocalization analyses presented in this study. Sharing of individual-level genome-wide SNP genotyping and DNAm profiling data is restricted by study-specific access policies. Interested researchers can contact the steering committees of the respective studies to inquire about access: 1. BASE-II, 2. BBHI, 3. LCBC. Requested data will be made available pending appropriate institutional data protection security measures and ethical approval of the requestor’s institution.

## Supplementary Tables

Can be found in Supplementary Tables document.

## Supplementary Figures

**Figure S1**. Chessboard plots for study-wide significant SNP-CpG association results in buccal mucosa. A. SNP-CpG pairwise associations in the BASE-II dataset (n=837). B. Pairwise SNP-CpG associations in the combined BBHI-LCBC dataset (n=690). Each dot represents a SNP-CpG pair that has exceeded the study-wide significance level (P<10^-14^; Methods). CpG positions are shown on the x-axis, and SNP positions are shown on the y-axis. CpG position and CpG density (#CpGs/Mb) are provided on the x axis, while SNP position and SNP density (#CpGs/Mb) are provided on the y axis. SNP-CpG pairs are coded according to their genomic distance: *cis* = for pairs within 1 Mb (green markers; appear as a diagonal line); long-range *cis* = for pairs on the same chromosome but >1Mb apart (purple markers); *trans* = for pairs located on different chromosomes (black markers).

**Figure S2**. Manhattan plots of *trans* acting SNP-CpG associations. A. BASE-II-buccal dataset (n=837). B. BBHI-LCBC-buccal (n=690) dataset. Each dot represents a SNP marker. Red dots indicate the top 1% SNPs across all detected *trans* associations.

**Figure S3**. Proportion of significant SNP-CpG pairs that replicate in three out of three or two out of three tissue types, or that are present only in one tissue. Results are separated depending on distance between CpG and SNP into A. *cis* regions, B. lr-*cis* regions, and C. *trans* regions.

**Figure S4**. Manhattan plots illustrating association results from A. AD GWAS, SMR results for an association between AD risk and DNAm in B) blood, C) buccals, and D) saliva. The red horizontal lines represent the genome-wide significance level for GWAS (P=5 × 10^−8^) and SMR analysis (P=1.6 × 10^-7^). Effects that passed the HEIDI test for heterogeneity (P≥0.05) are highlighted in red.

**Figure S5**. Manhattan plots illustrating association results from A. Parkinson’s disease GWAS, B. SMR results for an association between DNAm in blood dataset and Parkinson’s disease, C. SMR results for an association between DNAm in buccal dataset and Parkinson’s disease, D. SMR results for an association between DNAm in saliva dataset and Parkinson’s disease. The red horizontal lines represent the genome-wide significance level for GWAS (P=5 × 10^−8^) and SMR analysis (P=1.6 × 10^-7^). Effects that passed the HEIDI test for heterogeneity (P≥0.05) are highlighted in red.

**Figure S6**. Flowchart of MR study design and analysis strategies applied. Each CpG is annotated with a gene from the Illumina manifest. CpGs in intergenic regions are assumed to be one independent genomic locus each.

**Figure S7**. Forest plot of MR results for association between AD risk and DNAm using four different MR methods for A) blood and B) buccal tissue (no study-wide significant results were observed for saliva). The plot shows study-wide significant (p<1.47×10^-4^) results of MR analyses using at least three independent (r^2^<0.01, 1000 kb) SNPs as instrumental variables.

**Figure S8**. Forest plot of MR results for association between Parkinson’s disease risk and DNAm using four different MR methods for A) blood, B) buccal, and C) saliva tissue. The plot shows study-wide significant (p<1.47×10^-4^) results of MR analyses using at least three independent (r^2^<0.01, 1000 kb) SNPs as instrumental variables.

