## Supplementary figures and images for "Genome-wide QTL mapping across three tissues highlights several Alzheimer’s and Parkinson’s disease loci potentially acting via DNA methylation"

### Supplementary Figure S1A

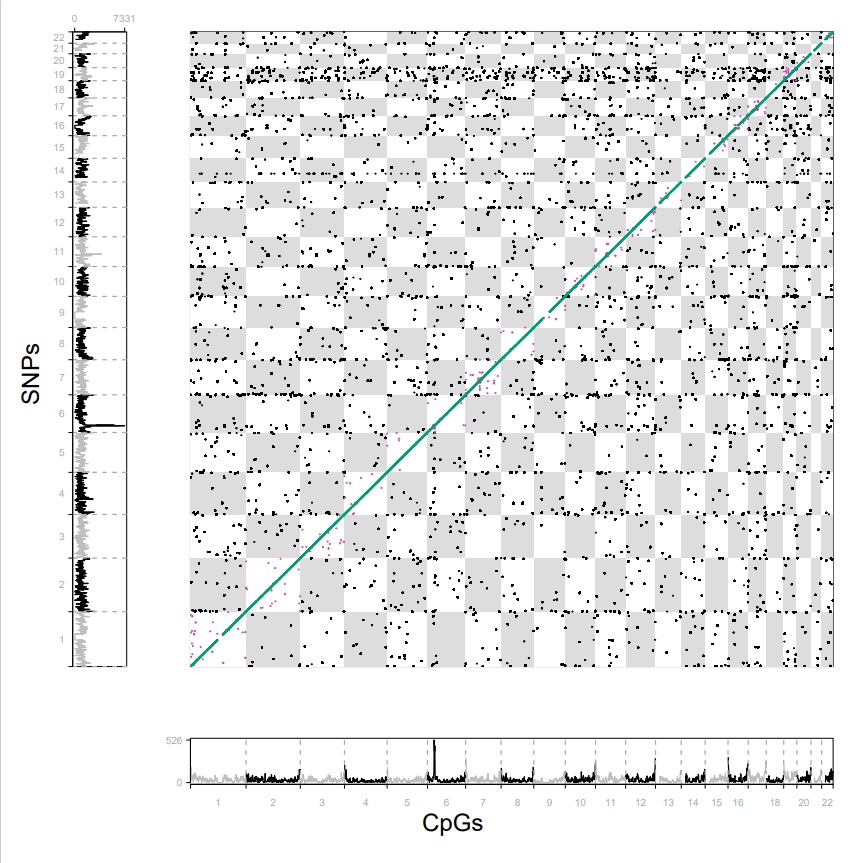

### Supplementary Figure S1B

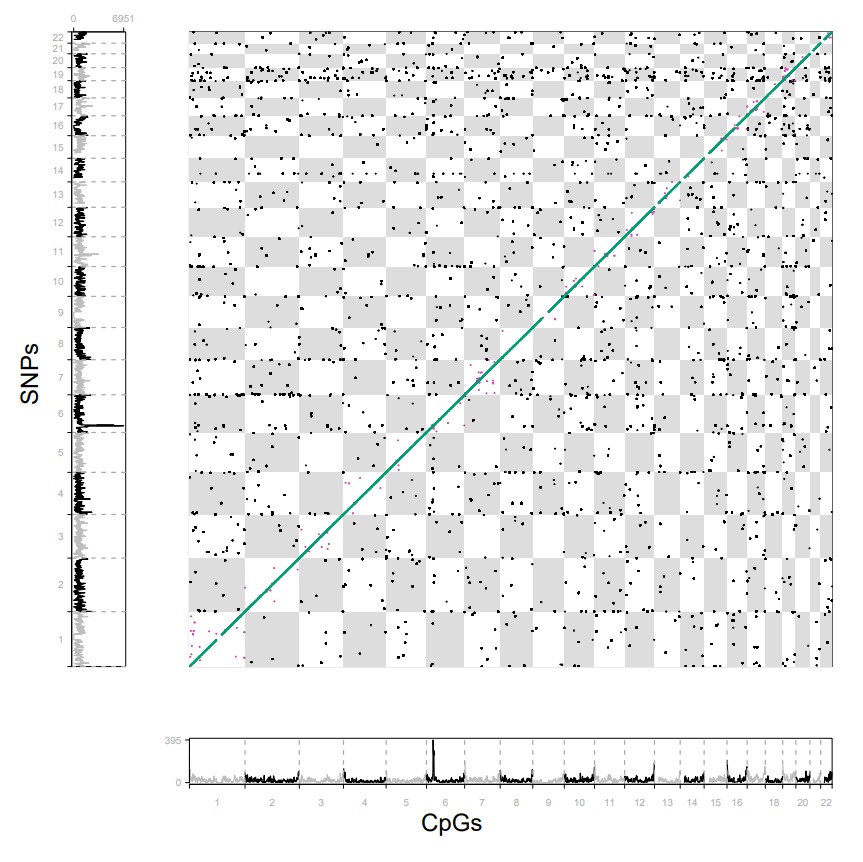

### Supplementary Figure S2A

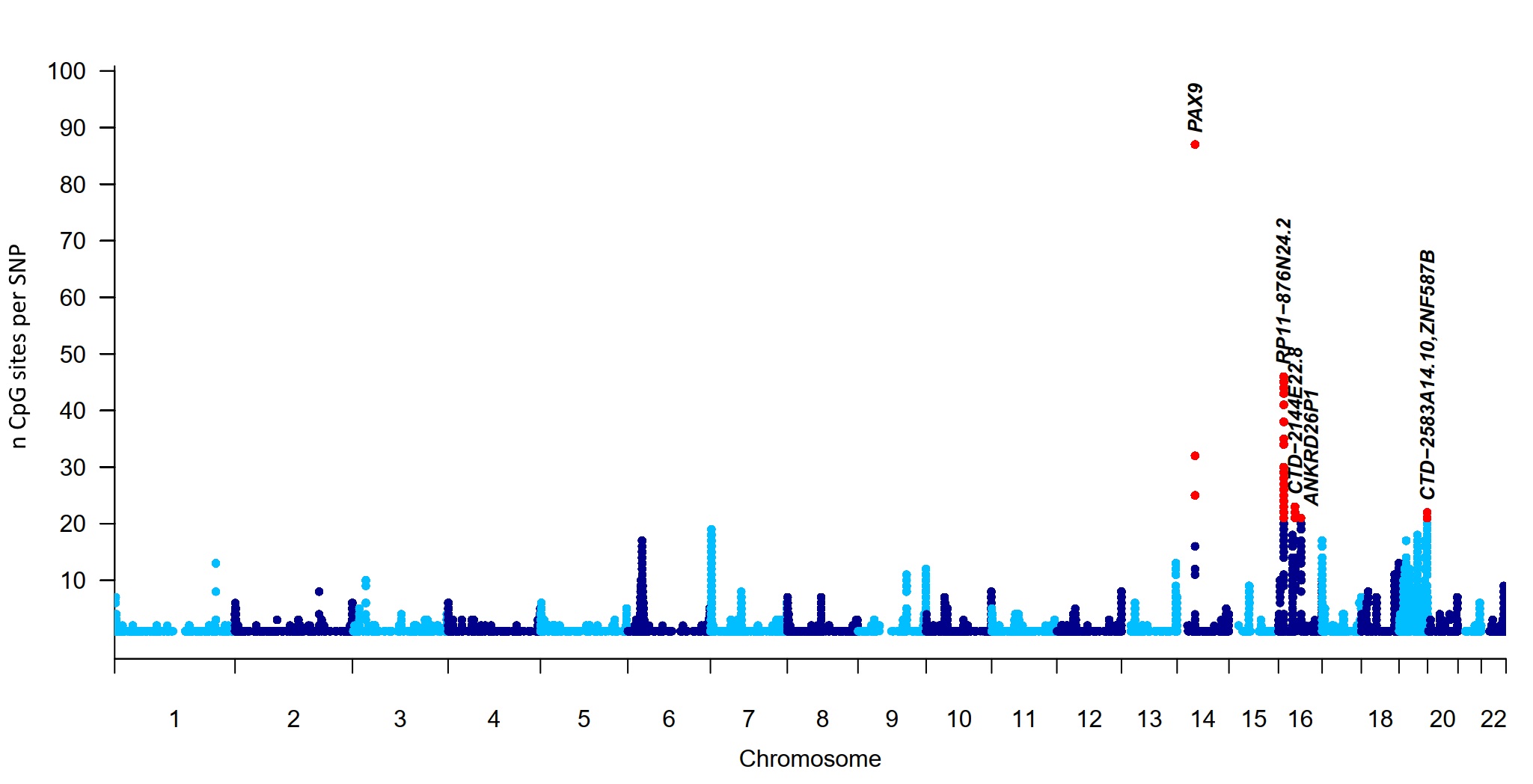

### Supplementary Figure S2B

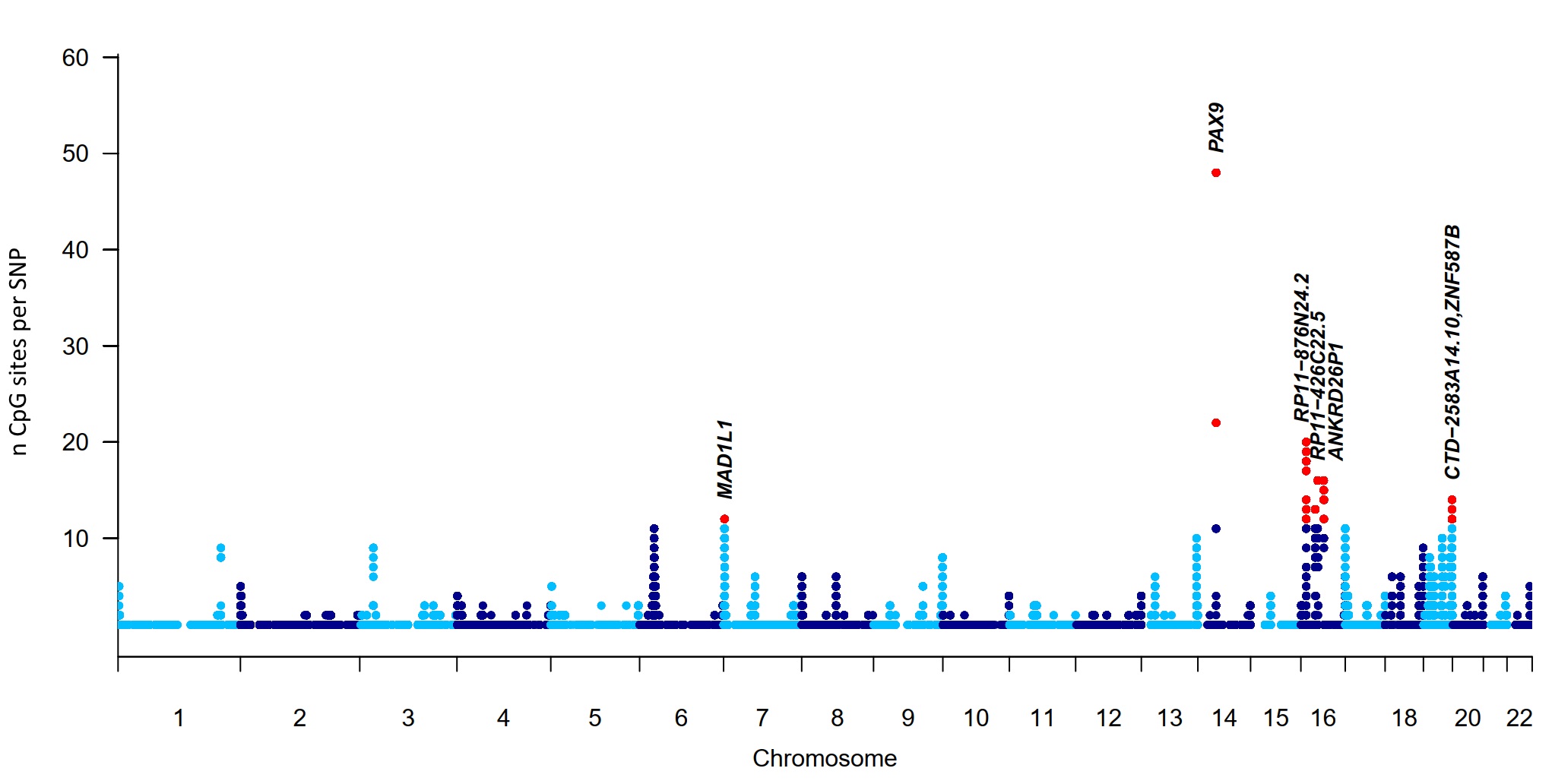

### Supplementary Figure S3

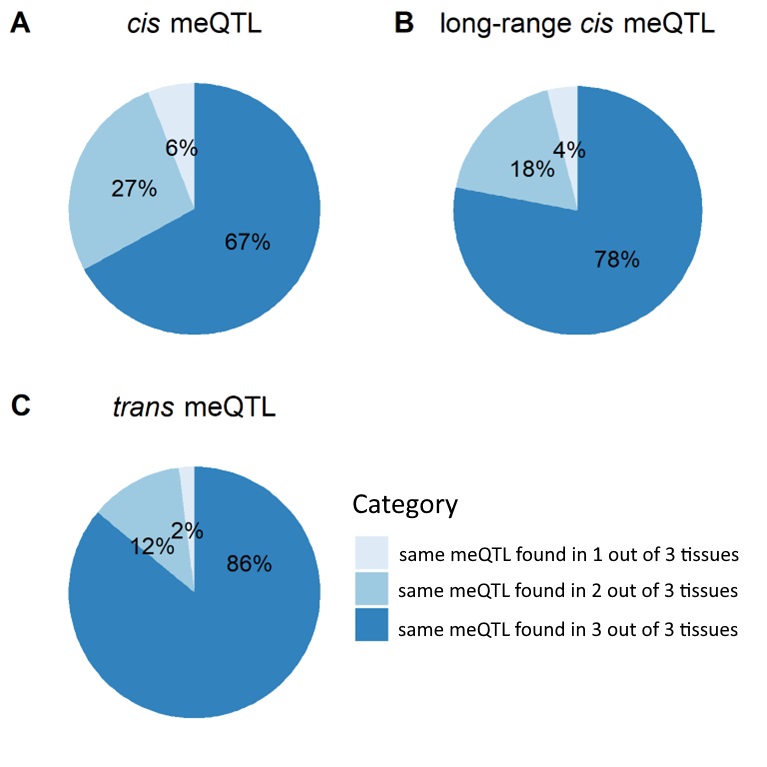

### Supplementary Figure S4

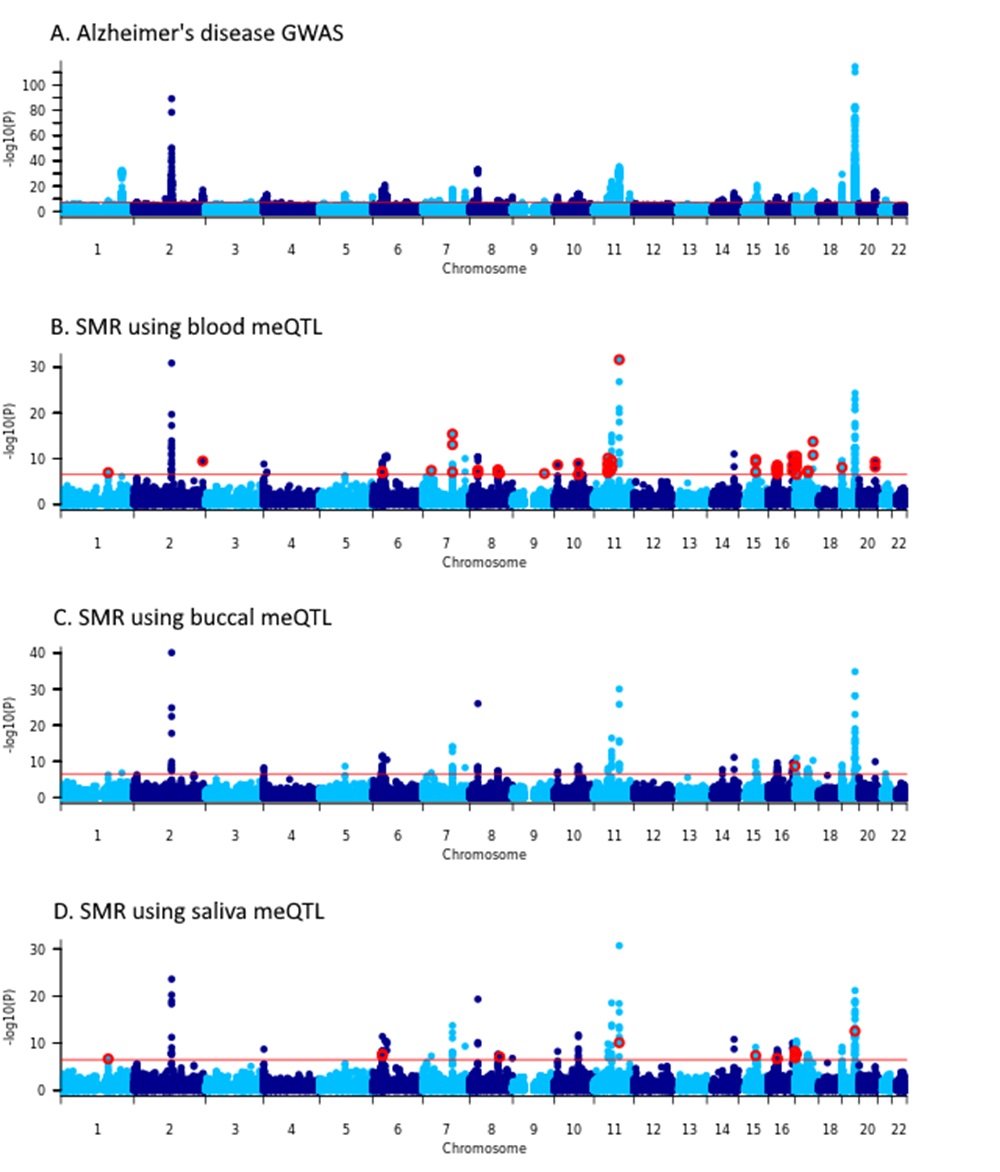

### Supplementary Figure S5

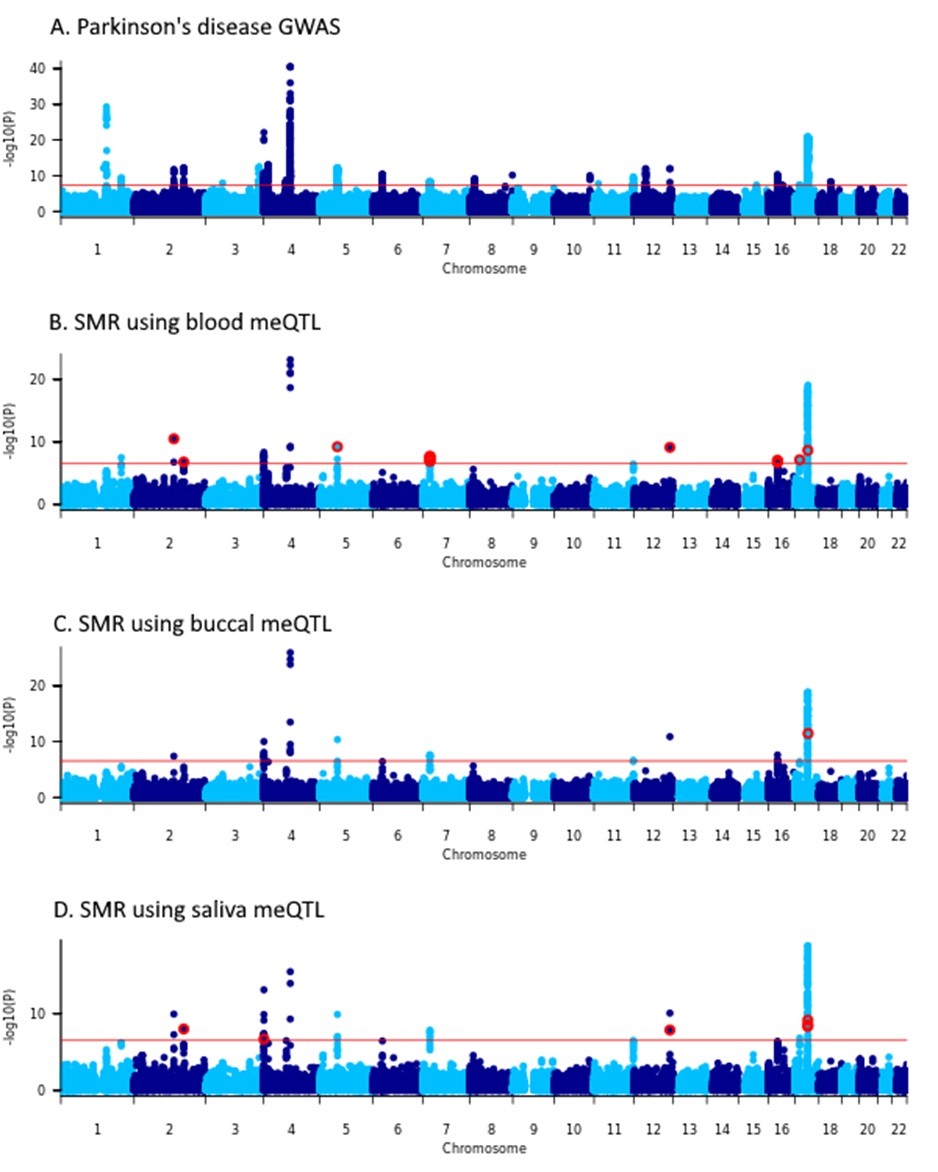

### Supplementary Figure S6

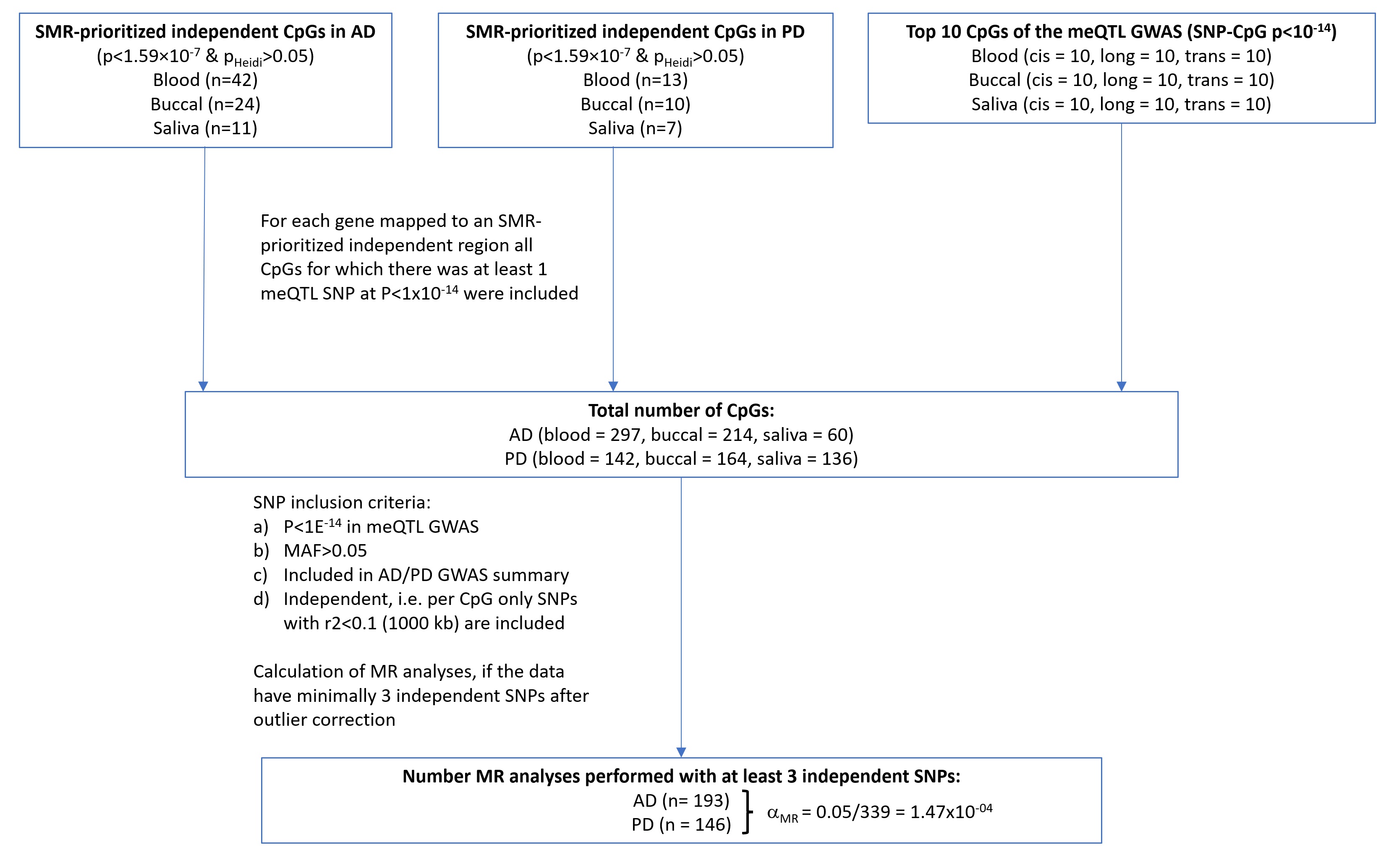

### Supplementary Figure S7A

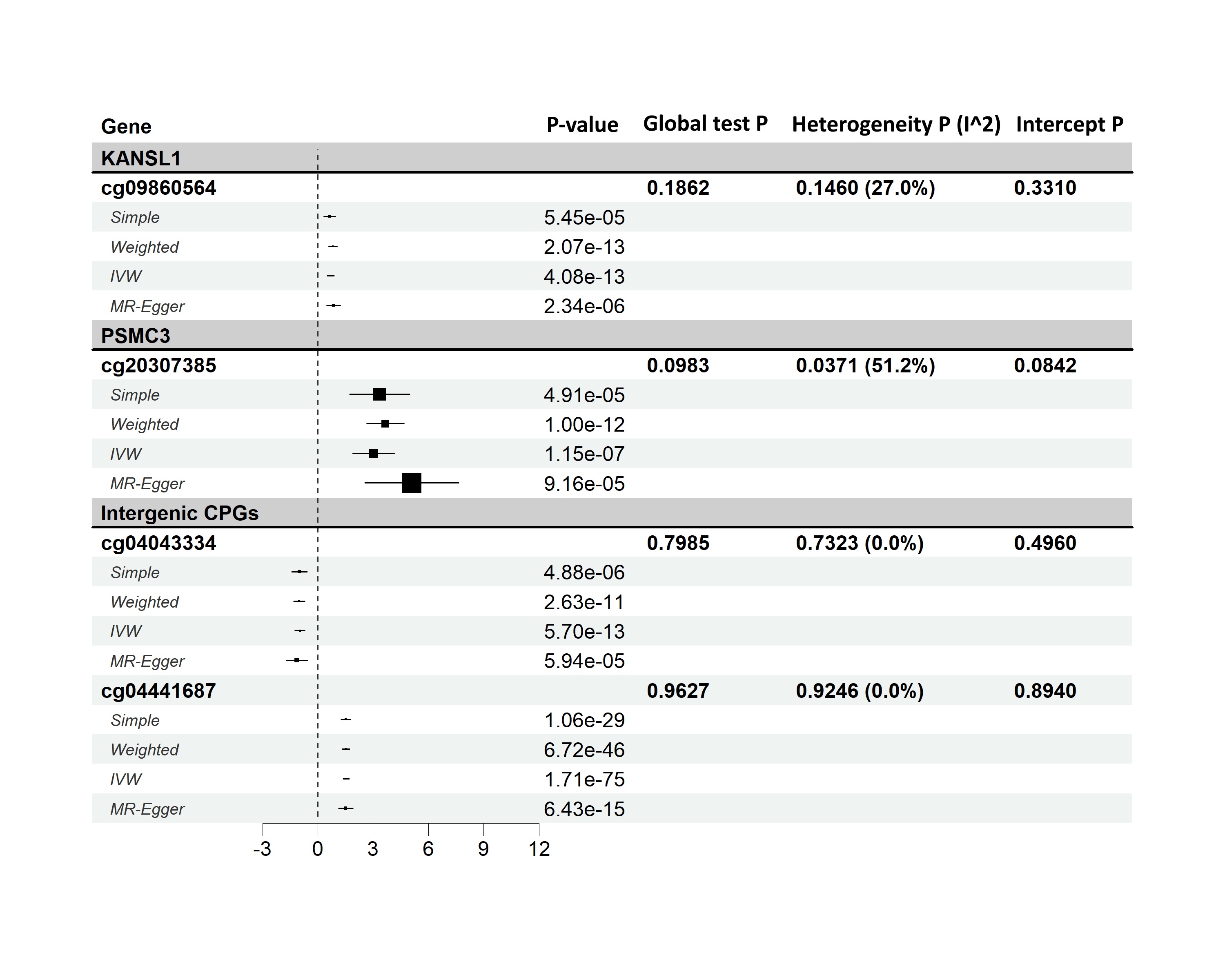

### Supplementary Figure S7B

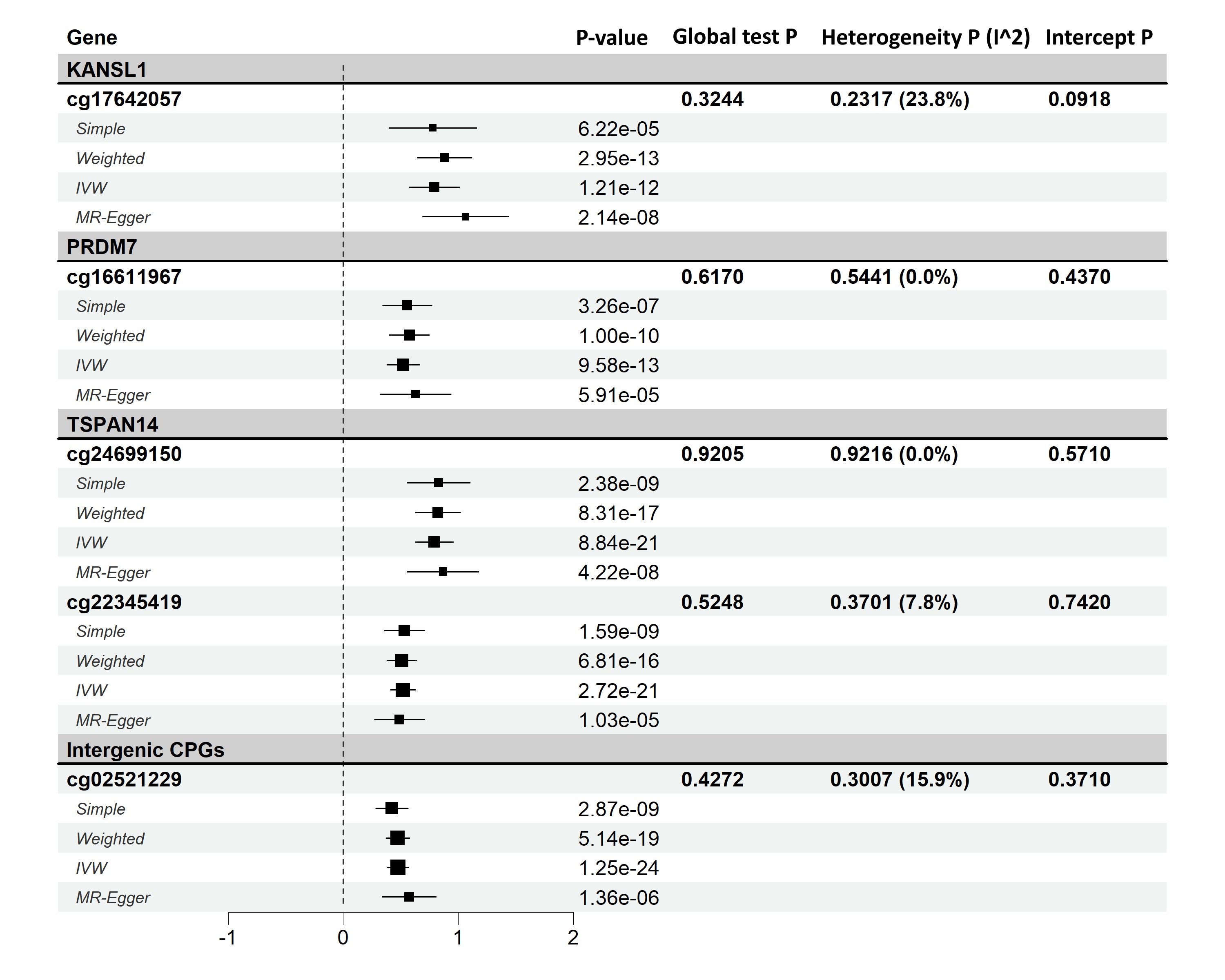

### Supplementary Figure S8A

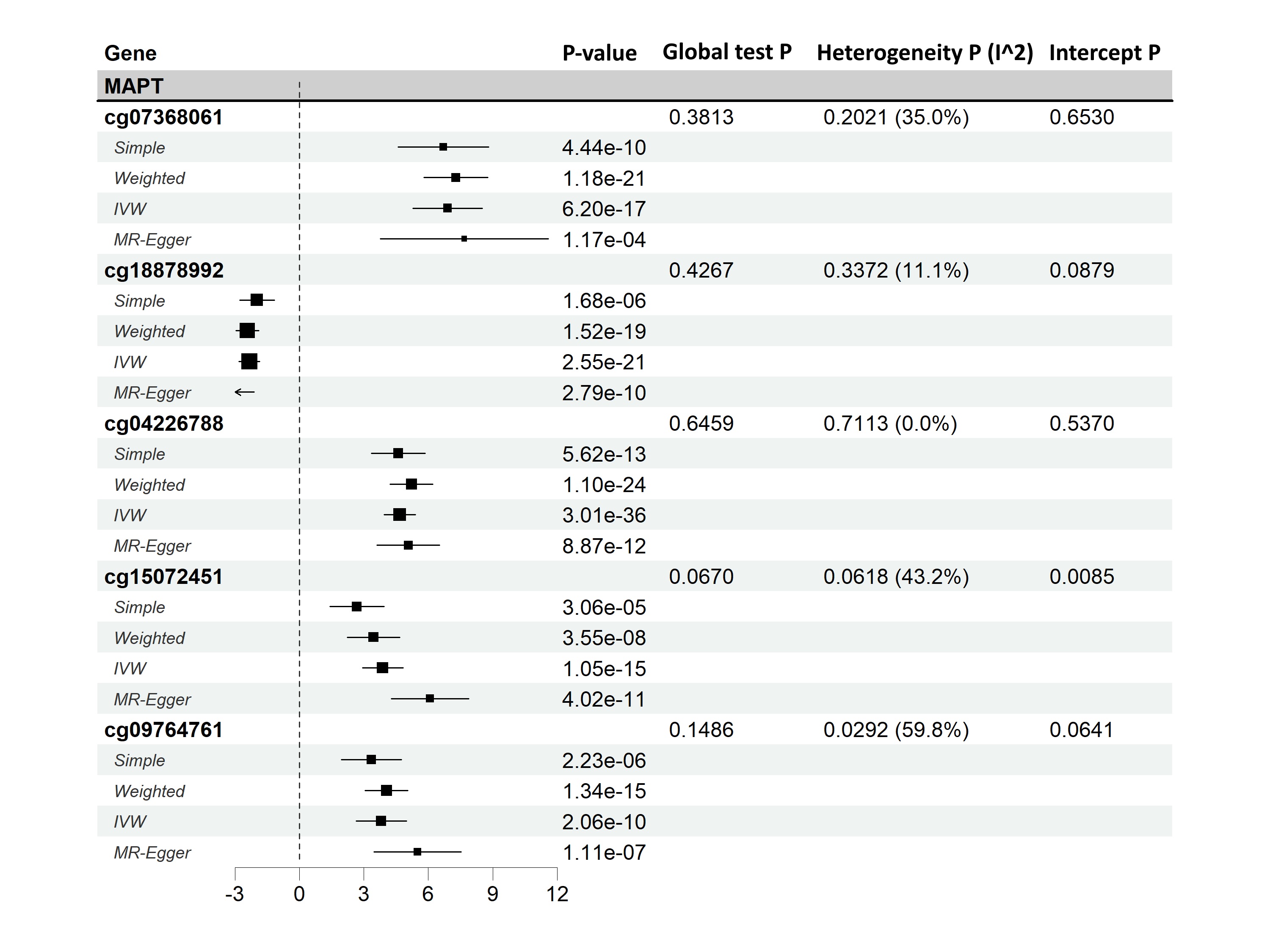

### Supplementary Figure S8B

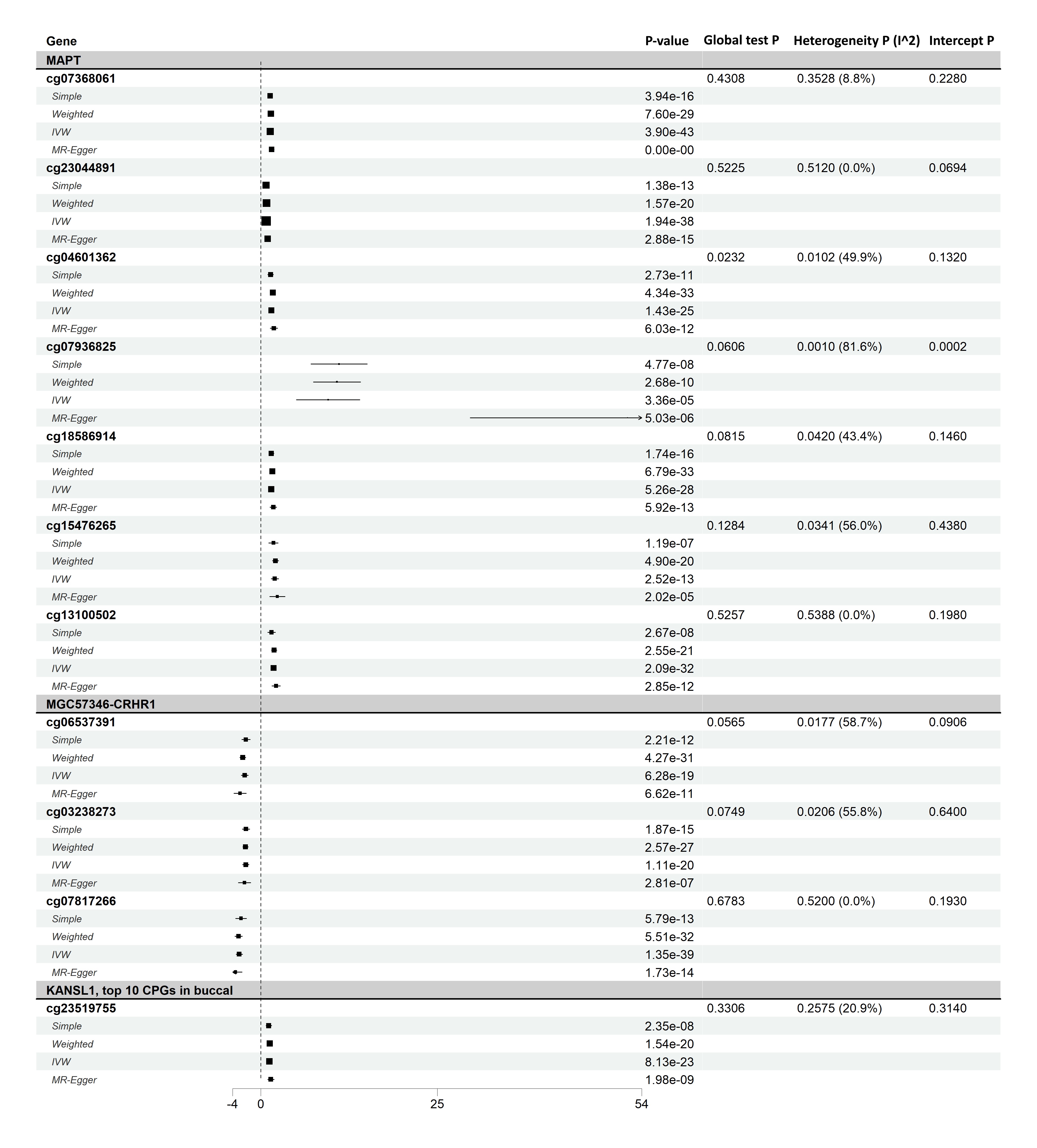

### Supplementary Figure S8C

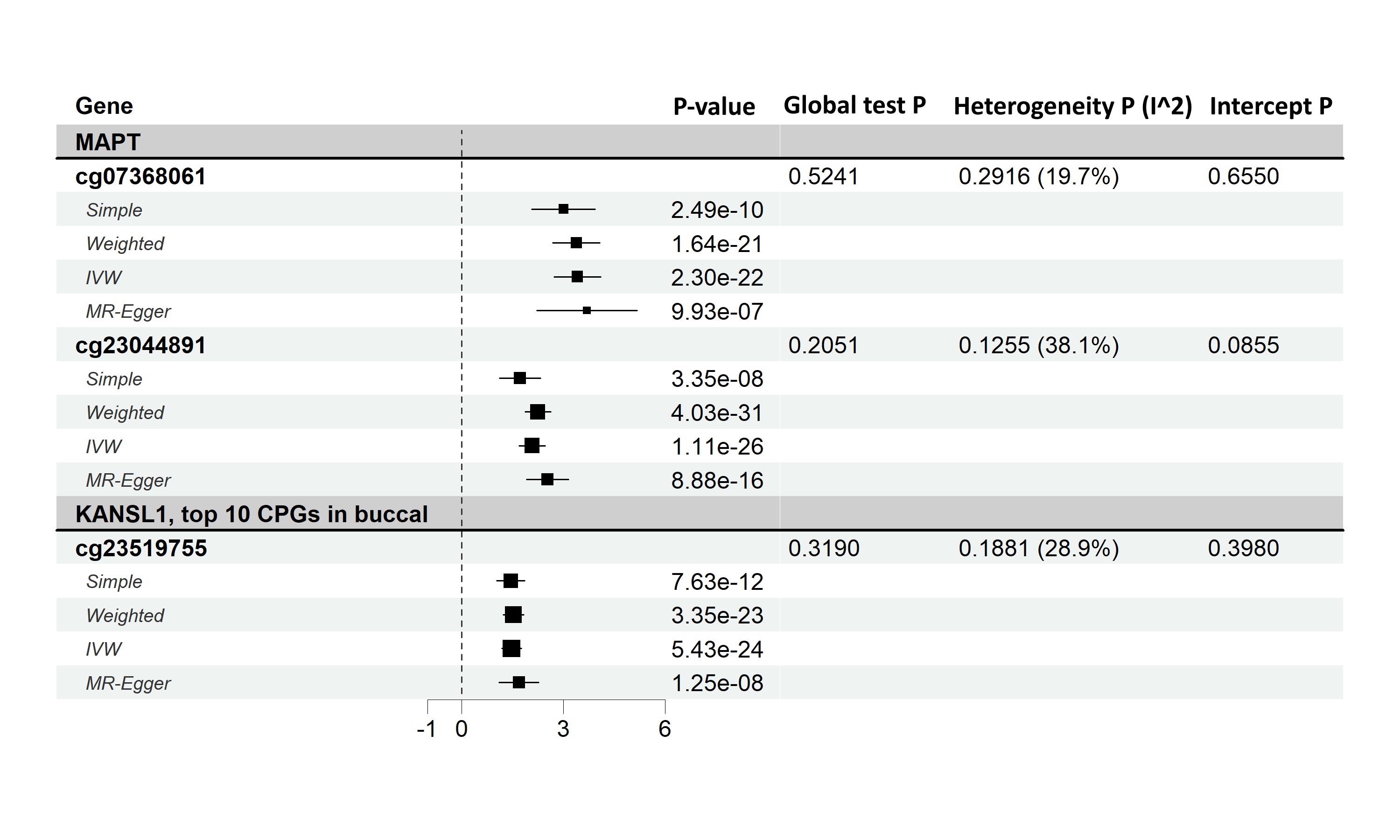
